# Epidemic waves of COVID-19 in Scotland: a genomic perspective on the impact of the introduction and relaxation of lockdown on SARS-CoV-2

**DOI:** 10.1101/2021.01.08.20248677

**Authors:** Samantha J Lycett, Joseph Hughes, Martin P McHugh, Ana da Silva Filipe, Rebecca Dewar, Lu Lu, Thomas Doherty, Amy Shepherd, Rhys Inward, Gianluigi Rossi, Daniel Balaz, Rowland R Kao, Stefan Rooke, Seb Cotton, Michael D Gallagher, Carlos Balcazar Lopez, Áine O’Toole, Emily Scher, Verity Hill, John T McCrone, Rachel M Colquhoun, Ben Jackson, Thomas C Williams, Kathleen A Williamson, Natasha Johnson, Katherine Smollett, Daniel Mair, Stephen Carmichael, Lily Tong, Jenna Nichols, Kirstyn Brunker, James G Shepherd, Kathy Li, Elihu Aranday-Cortes, Yasmin A Parr, Alice Broos, Kyriaki Nomikou, Sarah E McDonald, Marc Niebel, Patawee Asamaphan, Igor Starinskij, Natasha Jesudason, Rajiv Shah, Vattipally B Sreenu, Tom Stanton, Sharif Shaaban, Alasdair MacLean, The COVID-19 Genomics UK (COG-UK) consortium, Mark Woolhouse, Rory Gunson, Kate Templeton, Emma C Thomson, Andrew Rambaut, Matthew T.G. Holden, David L Robertson

## Abstract

The second SARS virus, SARS-CoV-2, emerged in December 2019, and within a month was globally distributed. It was first introduced into Scotland in February 2020 associated with returning travellers and visitors. By March it was circulating in communities across the UK, and to control COVID-19 cases, and prevent overwhelming of the National Health Service (NHS), a ‘lockdown’ was introduced on 23rd March 2020 with a restriction of people’s movements. To augment the public health efforts a large-scale genome epidemiology effort (as part of the COVID-19 Genomics UK (COG-UK) consortium) resulted in the sequencing of over 5000 SARS-CoV-2 genomes by 18th August 2020 from Scottish cases, about a quarter of the estimated number of cases at that time. Here we quantify the geographical origins of the first wave introductions into Scotland from abroad and other UK regions, the spread of these SARS-CoV-2 lineages to different regions within Scotland (defined at the level of NHS Health Board) and the effect of lockdown on virus ‘success’. We estimate that approximately 300 introductions seeded lineages in Scotland, with around 25% of these lineages composed of more than five viruses, but by June circulating lineages were reduced to low levels, in line with low numbers of recorded positive cases. Lockdown was, thus, associated with a dramatic reduction in infection numbers and the extinguishing of most virus lineages. Unfortunately since the summer cases have been rising in Scotland in a second wave, with >1000 people testing positive on a daily basis, and hospitalisation of COVID-19 cases on the rise again. Examining the available Scottish genome data from the second wave, and comparing it to the first wave, we find that while some UK lineages have persisted through the summer, the majority of lineages responsible for the second wave are new introductions from outside of Scotland and many from outside of the UK. This indicates that, while lockdown in Scotland is directly linked with the first wave case numbers being brought under control, travel-associated imports (mostly from Europe or other parts of the UK) following the easing of lockdown are responsible for seeding the current epidemic population. This demonstrates that the impact of stringent public health measures can be compromised if following this, movements from regions of high to low prevalence are not minimised.

## Introduction

In 2002/03 a new human coronavirus, SARS-CoV, emerged from horseshoe bats in China. These outbreaks were controlled by public health interventions and the total number of infected people was held at about 8000, deaths to 774, and virus eradication achieved (Anderson et al. 2004). Subsequent surveillance efforts for SARS-like viruses identified a reservoir of *Severe acute respiratory syndrome-related coronaviruses* in horseshoe bats and warned of risks for future spill over (Li et al. 2005, Menachery et al. 2015). A virus of the same species, named SARS-CoV-2 (Gorbalenya et al. 2020), was detected in a cluster of infections in Wuhan city, China, in December 2019. The first COVID-19 death occurred on the 11th January and by the 24th January, there were >800 confirmed cases and 17 deaths recorded (Wang et al. 2020). A pandemic was declared by the WHO on the 12th March 2020 and Europe became the epi-centre of the SARS-CoV-2 pandemic.

As of November 2020 there have been >58 million cases recorded and >1.4 million deaths due to COVID-19. These numbers are despite the use of unprecedented non-pharmaceutical public health interventions to restrict virus transmission. For example, on the 23rd January the first ‘lockdown’ was imposed on several cities in Hubei province. While efforts to control spread in China were largely successful owing to strict use of control measures -- with reported deaths being below 5,000, 10 times fewer deaths than the UK despite a population 20 times larger (Burki 2020) -- the control of people’s movement on an unprecedented scale, coupled with damage to economies, has made lockdowns controversial. Here, by taking a genomics perspective, we analyse the response in the UK with a focus on Scotland.

Genome-based studies of the SARS-CoV-2 variants in different geographical regions identified multiple introductions associated with international travel routes (for example, (Bugembe et al. 2020, Tayoun et al. 2020, Worobey et al. 2020). The introductions to the UK (>1400 events) have been recorded to early February and throughout that month (20% of transmission lineages) with the majority (80%) being established in March (Plessis et al. 2020). By the end of March we have estimated there were at least 283 introductions of SARS-CoV-2 into Scotland, the first recorded case being the 28th February, and phylogenetic analysis suggests an earlier introduction around the 19th February (Filipe et al. 2020). As the proportion of infected individuals increased markedly, and without access to suitable treatments, it became clear health services would struggle to cope with those with severe disease and a UK stay-at-home lockdown began on the 23rd March.

As part of the public health response, the COVID-19 Genomics UK (COG-UK) consortium, was formed to sequence and analyse the genetic material of SARS-CoV-2 as it spread in the four regions: England, Northern Ireland, Scotland and Wales. The consortium applies genomic sequencing technologies to the UK epidemic and provides crucial information on the surveillance of the circulating variants and on virus spread, contributing to local epidemiological investigations, monitoring of hospital-associated transmissions and nosocomial infections, and the study of the evolution of the virus, for example, of variants of antigenic significance. In this paper we focus on the spread of the dominant SARS-CoV-2 lineages that were introduced into Scotland and the association between lockdown and their circulation. We compare these first wave (a period from February 2020 to the easing of restrictions in July 2020) lineages to the variants responsible for the second wave (a period from August 2020 onwards). The two main lessons to be learned are (i) the lockdown was associated with a highly effective reduction in infection numbers and virus diversity, and (ii) once the virus is brought under control, restricting movement into geographical regions with very low prevalence is important to prevent re-introductions and the overwhelming of public health efforts.

## Results

### The first wave -- diversity of lineages seen in Scotland

SARS-CoV-2 sequences can be classified into distinct lineages (Rambaut et al. 2020). Of the 164 named lineages in the global dataset 101 are present in the UK reflecting the broad diversity of the SARS-CoV2 introductions into the UK and confirming our previous analysis to the end of March 2020 (Filipe et al. 2020). Sequences classified as lineage B.1 and B.1.1 account for over 50% both globally and for the UK as a whole. Of the global diversity, a wide sample was introduced into Scotland (Supplementary figure 1), and representatives of 55 of the named global lineages are observed. 40% of the Scottish sequences were B.1 and B.1.1. Reflecting different travel patterns, there was a higher proportion of B.1.5 in Scotland than the rest of the UK (6% of Scotland sequences, <2% rest of UK); and global lineage B.1.93 was almost exclusive to Scotland (representing 14% of Scotland sequences, but only 12 sequences from the rest of the UK). Lineages with a greater total number of sequences in Scotland than the rest of UK, and for which there are significantly more sequences sampled (Fisher test, P<0.0001, and Bonferroni correction) are: A.2, A.3, A.5, B.1.1.12, B.1.1.14, B.1.1.20, B.1.1.28, B.1.100, B.1.101, B.1.40, B.1.5.10, B.1.69, B.1.70, B.1.71, B.1.77, B.1.89, B.1.9, B.1.90, B.1.93, B.16, presumably reflecting different travel destinations as we previously reported (Filipe et al. 2020).

This SARS-CoV-2 diversity circulating in Scotland is spread across the different regions of the country with variants from the global lineages intermingling in the different NHS Health Boards (Figure 1A), which represent the regional organisation of the NHS within Scotland. Importantly, the number of patient samples for which genome sequences have been obtained, 5137 genomes by the 18th August 2020, captures sufficiently (at about ∼25% of tested positive cases) the distribution of infections in Scotland since March (Figure 1Bl). It is important to appreciate that modelled prevalence estimates indicate that the number of recorded positive tests are only a fraction of the true number of cases.

**Figure 1.**
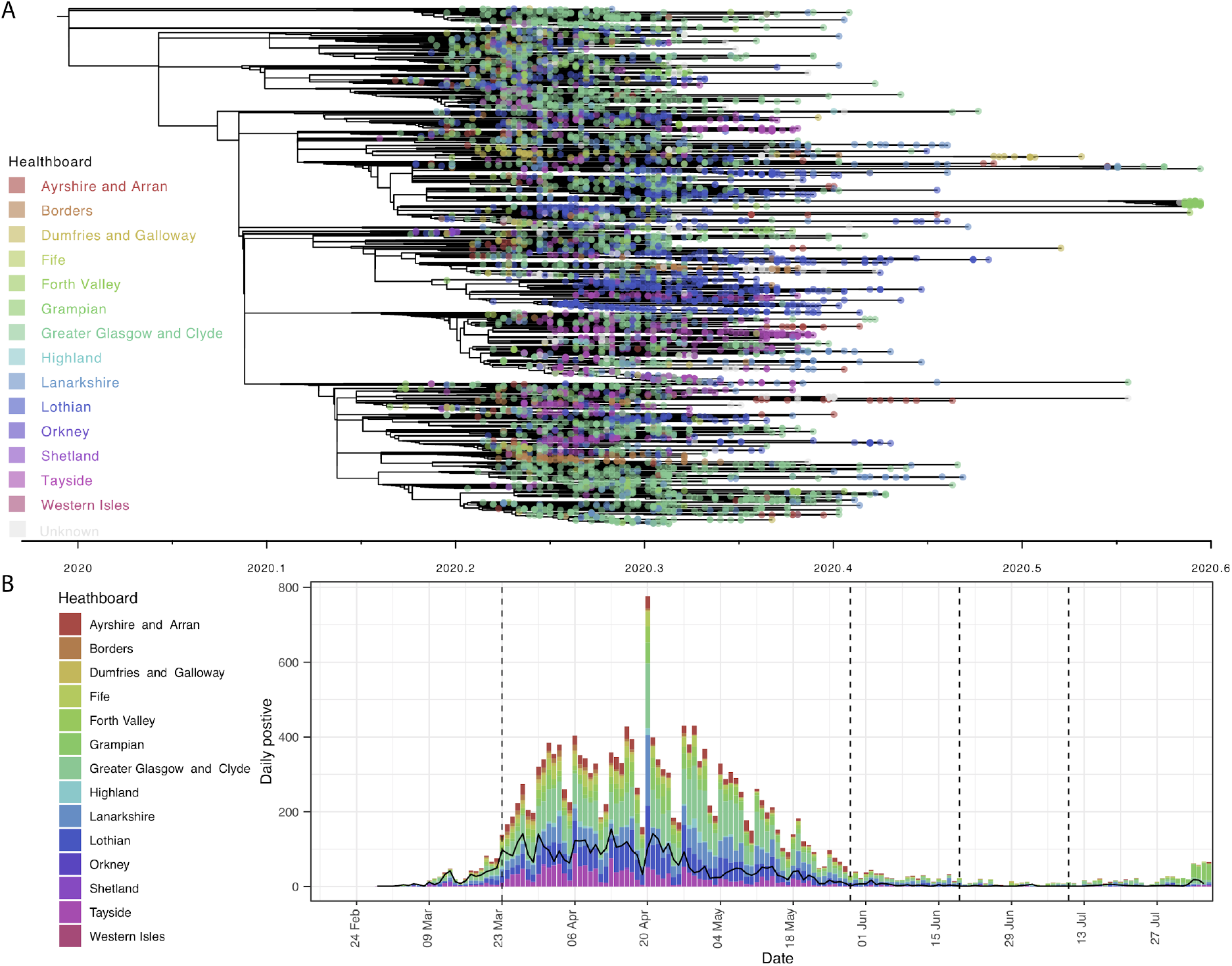
A| SARS-CoV-2 diversity circulating in Scotland up to 18th August 2020 represented by phylogenetic tree (inferred with TimeTree) with Scottish NHS Health Board focus (see key). B| the number of sequences (in black) versus the recorded positive cases by Scottish NHS Health Board plotted on the same time scale. The four vertical dashed lines represent the beginning of the lockdown (23rd March 2020) and its easing: movement to phase 1 (29th March 2020), phase 2 (19th June 2020) and phase 3 (10th July 2020).

A sampling strategy (see Methods) was followed to ensure representation of all of the regions of Scotland and relative frequencies of infections in the 14 Scottish NHS Health Boards (Figure 2). This high-coverage genome-level surveillance data demonstrates a clear rise and fall of COVID-19 in line with recorded cases (Figure 1B), and its resolution, among the highest in the world for a relatively small population (∼5.5 million people), permits a detailed characterisation of the spread of the virus within a single country. First, however, in order to track spread it is necessary to infer the distinct lineages associated with the introductions of SARS-CoV-2 into Scotland from other parts of the UK and globally.

**Figure 2.**
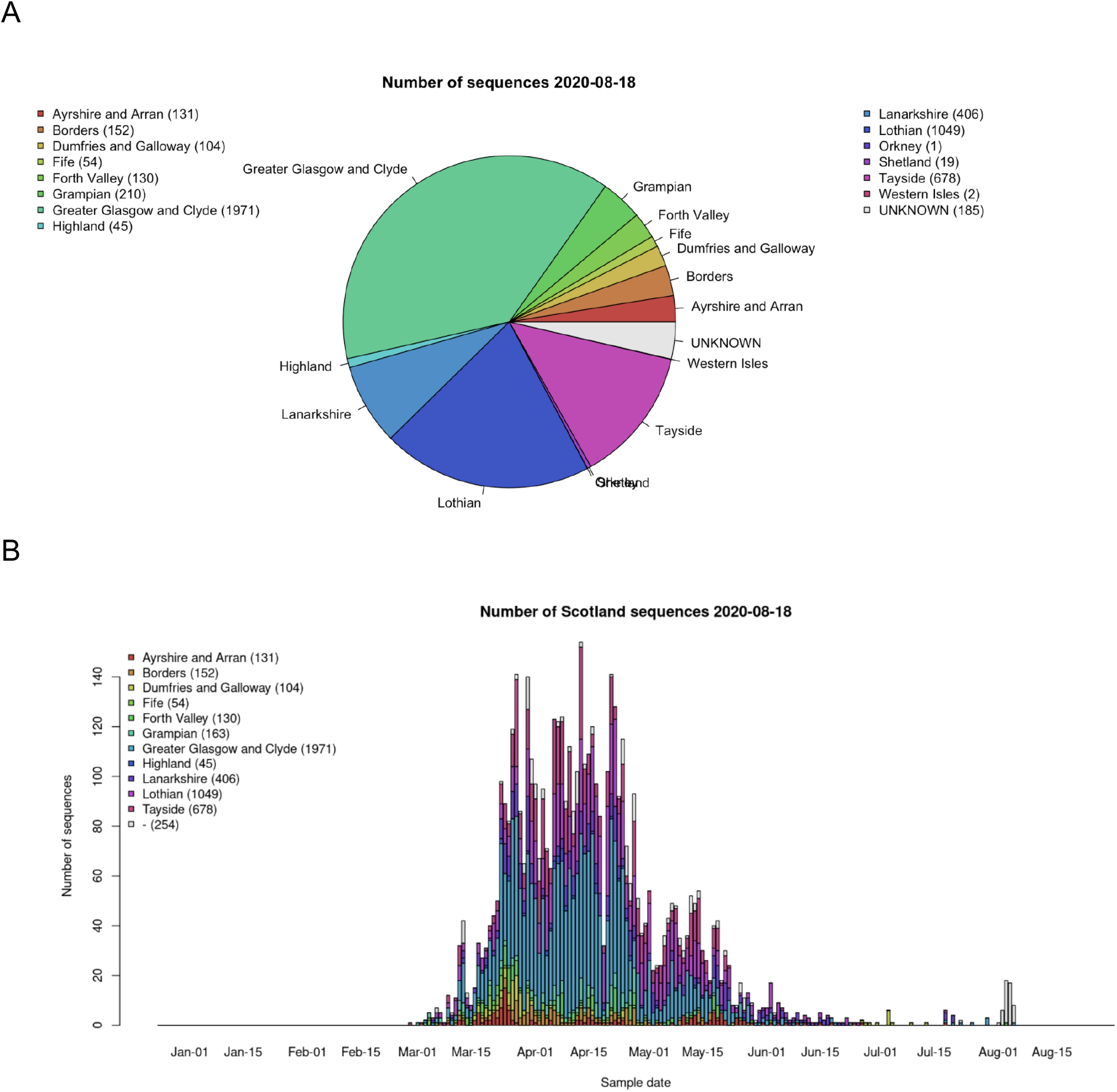
SARS-CoV-2 genome sequence counts by A| Scottish Health Board, and B| by sampling time from 28th February to 18th August 2020.

### The first wave -- estimates of introductions into Scotland pre-lockdown

Using a simple model to assign discrete locations to the ancestral nodes in the phylogeny (see Methods), the number of changes in the phylogeny from locations outside Scotland into Scotland were estimated. This gives an indication of the total number of imports, but is dependent on (and heavily biased by) the number of sequences sampled from different countries (some which are very low so unreliable) and the uncertainty and resolution in the phylogenetic tree. Using this approximate measure, we estimate 307 imports and 158 exports to and from Scotland respectively over the whole date range (Supplementary figure 2), close to our previous analysis (Filipe et al. 2020) where we estimated >283 introductions based on sequences sampled to the end March and known travel histories. With the caveat that sampling for non-UK regions can be sparse, most of these imports (50%) appear to come from England, 24% from the rest of Europe, and 7-8% each from Wales, Asia and North America (Supplementary figure 2A). The estimated proportion of the total exports that went back into England and Europe are lower than the imports (34% and 18% respectively), but the proportion is higher for Wales (9%), and North America (27%). The proportion of all exports from Scotland into mainland Europe (subject to some biases, see Methods) was approximately the same as the proportion of imports.

A similar approach to estimate the number of location changes in the phylogeny from non-UK to UK resulted in 1540 introduced lineages, of which 283 were found in Scotland. Of these 283 lineages, most (217) are represented by five variants or less (154 by only one), leaving 66 lineages represented by >five variants. These introductions to the UK were estimated from the global tree, and each introduction was given a ‘UK lineage number’ (see Methods). Some introductions gave rise to many subsequent infections in all parts of the UK, for example, the first sequences being sampled from places in England, and then subsequently sampled from places in Scotland. Other introduced lineages were first sampled in Scotland. Note, the number of introductions estimated from the number of lineages is not quite the same as the approximate import count above (Supplementary figure 2), because a lineage can be transmitted between the different UK regions on more than one occasion, e.g., England and Scotland, but only counted as imported to the UK once.

Using the sequences grouped into UK lineages, and also including other closely related sequences (from other UK lineages or sequences from outside the UK) in ‘extended’ lineages, we estimated time-scaled trees for each lineage/cluster. This permitted the inference of the ancestral locations of the internal nodes in the phylogeny (see Methods). Supplementary figure 2B summarises the start time of the UK lineages which have sequences in Scotland, along with their inferred ancestral locations, i.e., the time to most recent common ancestor of the sequences within the lineage within Scotland, and the location of their direct ancestral node. UK lineages in Scotland which have such direct ancestors estimated to be in Scotland are also included, e.g., when two Scottish lineages are detected, but they both go back to the same multifurcating ancestral node.

### The first wave -- distribution of diversity within Scotland over time

Looking at the different regions of Scotland (defined by NHS Health Boards) we can track both the diversity of the UK lineages circulating in a local region and how imports versus exports to other regions within Scotland change through time (Figure 3). This demonstrates how, once introduced, the virus spread rapidly within Scotland. Of the NHS Health Boards, NHS Glasgow and Greater Clyde has a large number of exports (and imports) to NHS Tayside and England pre-lockdown (Figure 4). This continues during lockdown and shows a high number of exports to NHS Lanarkshire with an increase in exports following lockdown. NHS Lothian (Figure 4) also shows a high number of exports to England (Supplementary figure 3 and http://sars2.cvr.gla.ac.uk/RiseFallScotCOVID/). On imports pre-lockdown NHS Greater Glasgow and Clyde has a relatively large number of imports from England (246) as does NHS Lothian (175), and this reduces to 44 and 45 respectively during lockdown. These estimates for other NHS Health Boards can be seen in Supplementary figure 3, with a summary of the number of imports and exports into each region.

**Figure 3.**
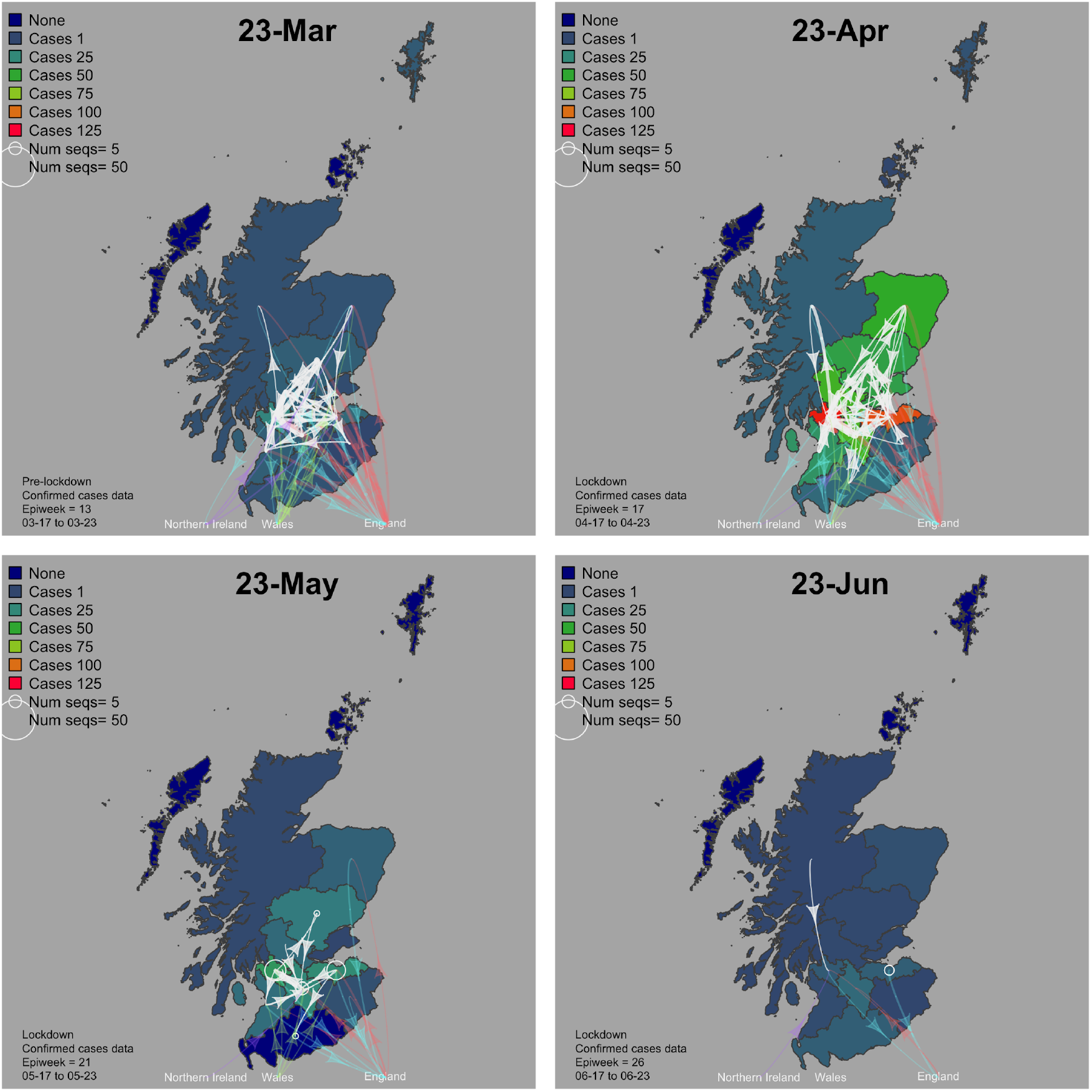
Maps showing the import and export relationships between the NHS Health Boards within Scotland at different timepoints from March to June. A time lapse video is available at http://sars2.cvr.gla.ac.uk/RiseFallScotCOVID/ Results for Orkney, Shetland and the Western Isles are not included as sequence numbers are too low for these regions.

**Figure 4.**
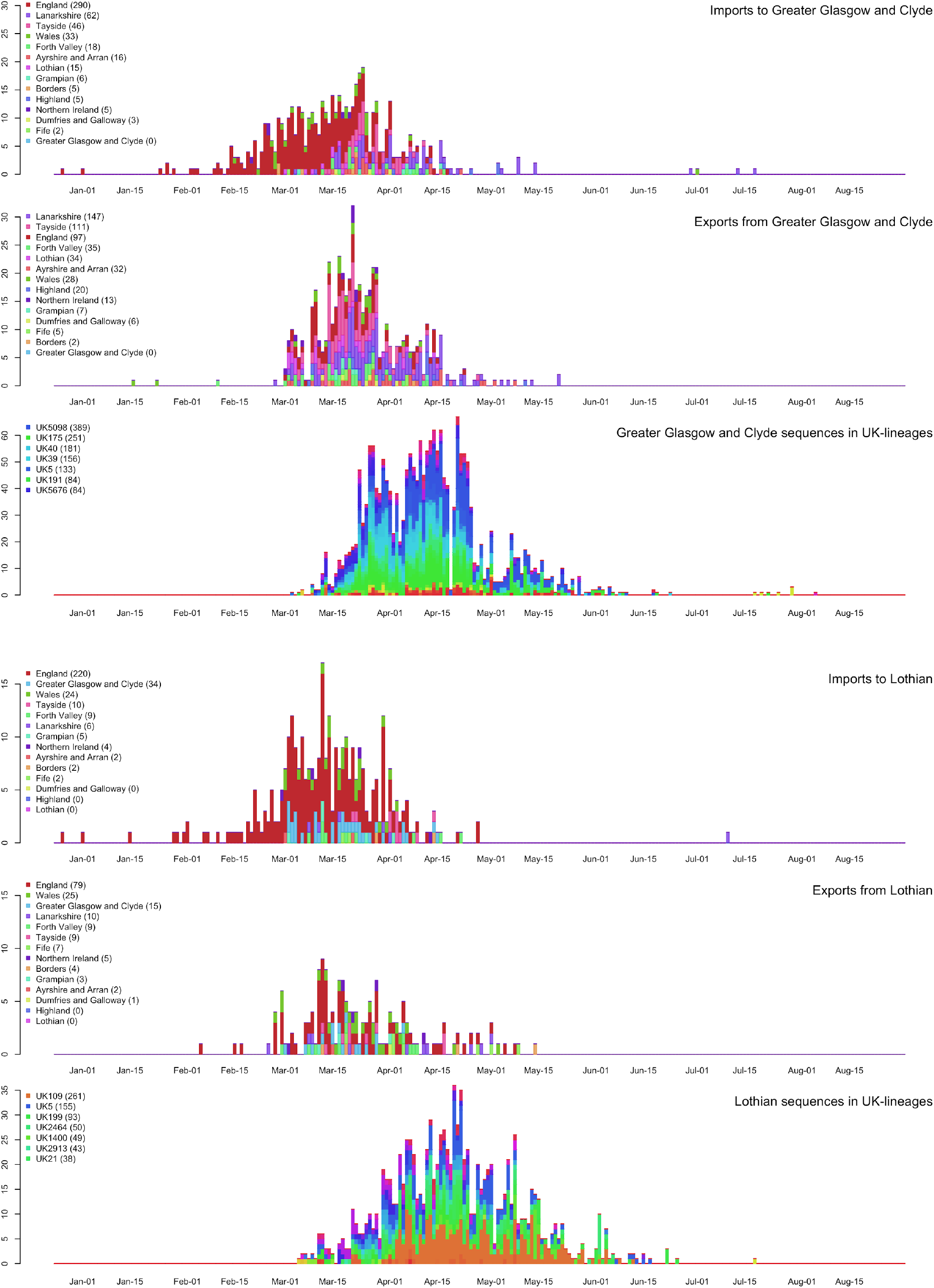
Timing of the imports and exports to Scottish NHS Health Board using NHS Greater Clyde and Glasgow and NHS Lothian as examples. See Supplementary figure 3 (and http://sars2.cvr.gla.ac.uk/RiseFallScotCOVID/) for other NHS Health Boards and a summary of the number of imports and exports into each region. The three plots for each NHS Health Board are as follows: top plot -- imports coloured by first NHS Health Board lineage detected in (rainbow colours) or England (red), Wales (green), Northern Ireland (purple). Note that the England (red) is a major source for both NHS Greater Glasgow and Clyde and Lothian in March; second row plot -- exports (same colouring as imports); third row plot -- number of sequences per NHS Health Board coloured by UK lineage number (other rainbow colours).

### The effect of lockdown

Although there was a lag from the start of the epidemic and the time the lockdown was imposed on the 23rd March, it can be directly linked with a substantial reduction in the number of COVID-19 cases, circulating SARS-CoV-2 variants and diversity in Scotland (Figure 4 and see Supplementary figures 3 and 4). The majority of lineages circulating decreased markedly and limited intra-country spread is observed by June (Figure 3). Interestingly for larger lineages there is a lag in lineage extinction with the time between observation of variants increasing steadily. The phylogenetic diversity in each NHS Health Board can be tracked through time (Supplementary figure 4). It can be seen that lockdown is associated with an immediate change in the diversity in the less populous NHS Health Boards but, not unexpectedly, a more delayed change in more populous regions, for example, in NHS Greater Glasgow and Clyde, NHS Lothian and NHS Lanarkshire the diversity only started decreasing 3-4 weeks after lockdown. However, that lockdown coincided with the elimination the majority of community transmissions in Scotland is clear from the data.

### Lineage dynamics in Scotland

SARS-CoV-2 variants enter and spread within the UK and Scotland, forming clusters and complex transmission chains. These chains or even whole lineages may die out due to stochastic epidemiological processes (even if R0>1, although the probability is lower than for R0<1), or due to the success of non-pharmaceutical restrictions in reducing transmission. Figure 5 (top) shows the number of sequences from Scotland available up until 21st October 2020, coloured by the UK lineage number, and Figure 5 (bottom) a heat map of the number of sequences from Scotland arranged into lineages – each horizontal line is a lineage, the colour at each epi-week representing the number of sequences. This shows there were many lineages which have sequences in Scotland before mid-July but these die out (388 of which 66 with size > 5 before epi-week 30), being replaced by lineages associated with new introductions by the end of August (epi-week 36 starts on 30th August 2020). From the end of August onwards, there were 30 new lineages with sequences in Scotland although only one had more than five sequences.

**Figure 5.**
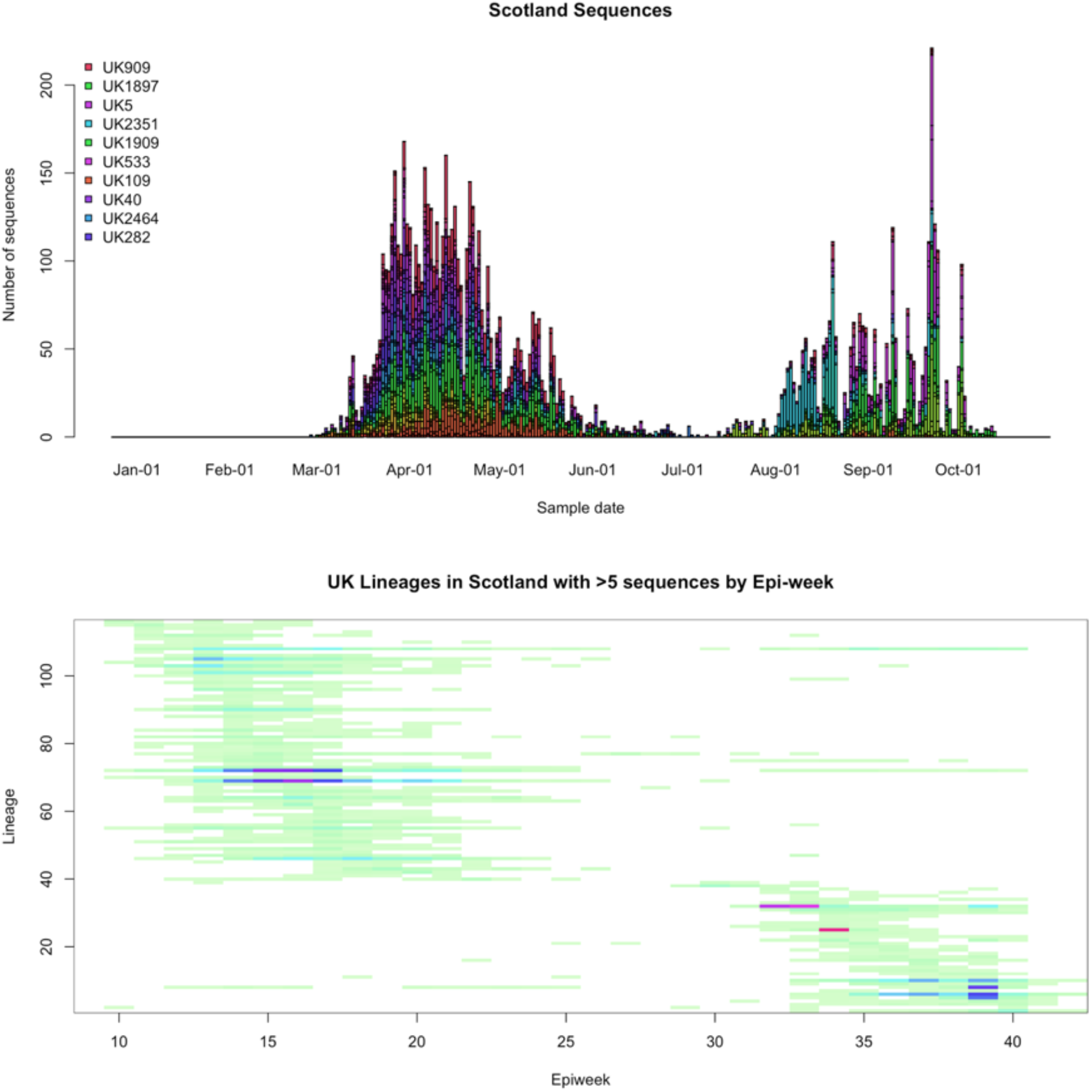
Re-emergence of SARS-CoV-2. UK lineages circulating in Scotland in first and second waves. Top | Number of sequences from Scotland, coloured by UK lineage number. Bottom | Heat map of sequences from Scotland in lineages by epi-week (minimum value = 1: pale green; maximum value = 169: bright magenta/red)

### The second wave -- persisting UK lineages in Scotland

There were 13 UK lineages which have sequences in Scotland (nine with more than five sequences) detected before epi-week 30 and also after epi-week 36 (see Supplementary table 1 for details). Bar plots of the number of sequences in England, Wales, Scotland and Northern Ireland with these UK lineages per epi-week are in the Supplementary figure 8. These Supplementary figures show firstly that the number of sequences in Scotland in these persisting lineages decrease or go to zero between epi-week 30 and 36. Since the number of sequences in Scotland represent a fraction of the true number of cases (presumably untested individuals with mild or no symptoms), it is important to note these persisting lineages may not have completely died out in Scotland. Secondly, however, the supplementary figures show that sequences from England dominate in these persisting lineages, although there are sequences from Wales in most of them, and from Northern Ireland in a minority – and there were no persisting lineages which contained only sequences from Scotland. Thirdly, the total number of sequences in the persisting lineages from anywhere in the UK ranged from five (UK650) to 13085 (UK5), with four lineages having at least 1000 sequences.

### The second wave -- origins and timing of new lineages

Following the easing of lockdown since July, and despite the continued implementation of physical distancing, community testing and contact tracing, the number of cases and associated lineages of SARS-CoV-2 has steadily increased (Figure 5). Without access to genome sequence data the origins of the second wave cases would be impossible to assess. However, using the 8603 sequences available by the 21st Oct 2020 we can compare these variants to those present in the first wave (Figure 5). This analysis shows that most of the latter are novel lineages, rather than persisting wave one lineages (Figure 5, bottom). Importantly, while some of the wave 2 lineages are re-introductions from other parts of the UK, the majority are introductions from outside of the UK which seed new lineages (Figure 6). Once introduced these lineages have grown within Scotland spreading to different regions (Figure 7).

**Figure 6.**
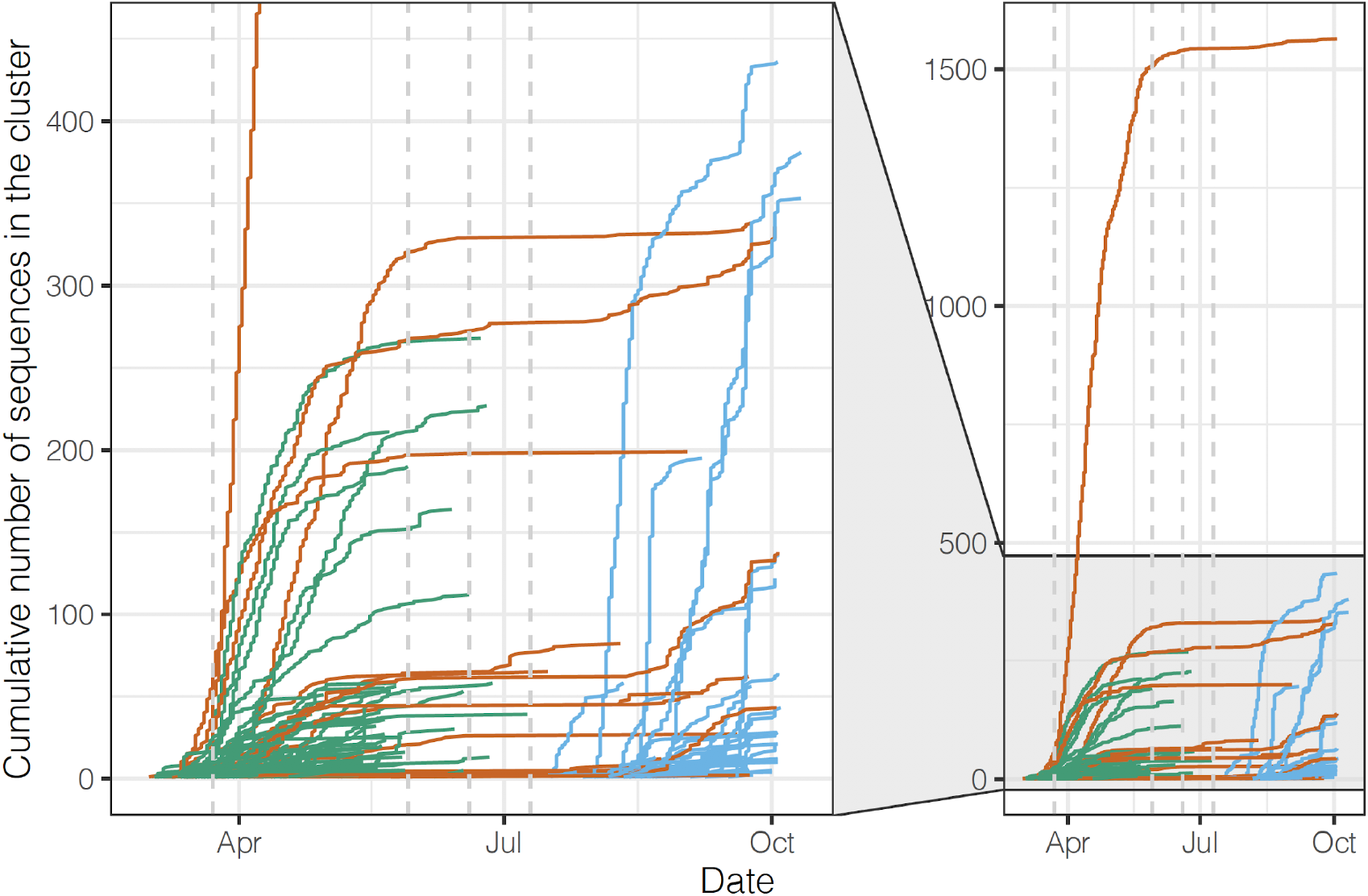
Cumulative Plot of the UK lineages present in Scotland through time depicting lineages that went extinct over the summer (green), persistent lineages (orange) and wave 2 lineages (blue). The four vertical dashed lines represent the beginning of the lockdown (23rd March 2020) and its easing: movement to phase 1 (29th March 2020), phase 2 (19th June 2020) and phase 3 (10th July 2020).

**Figure 7.**
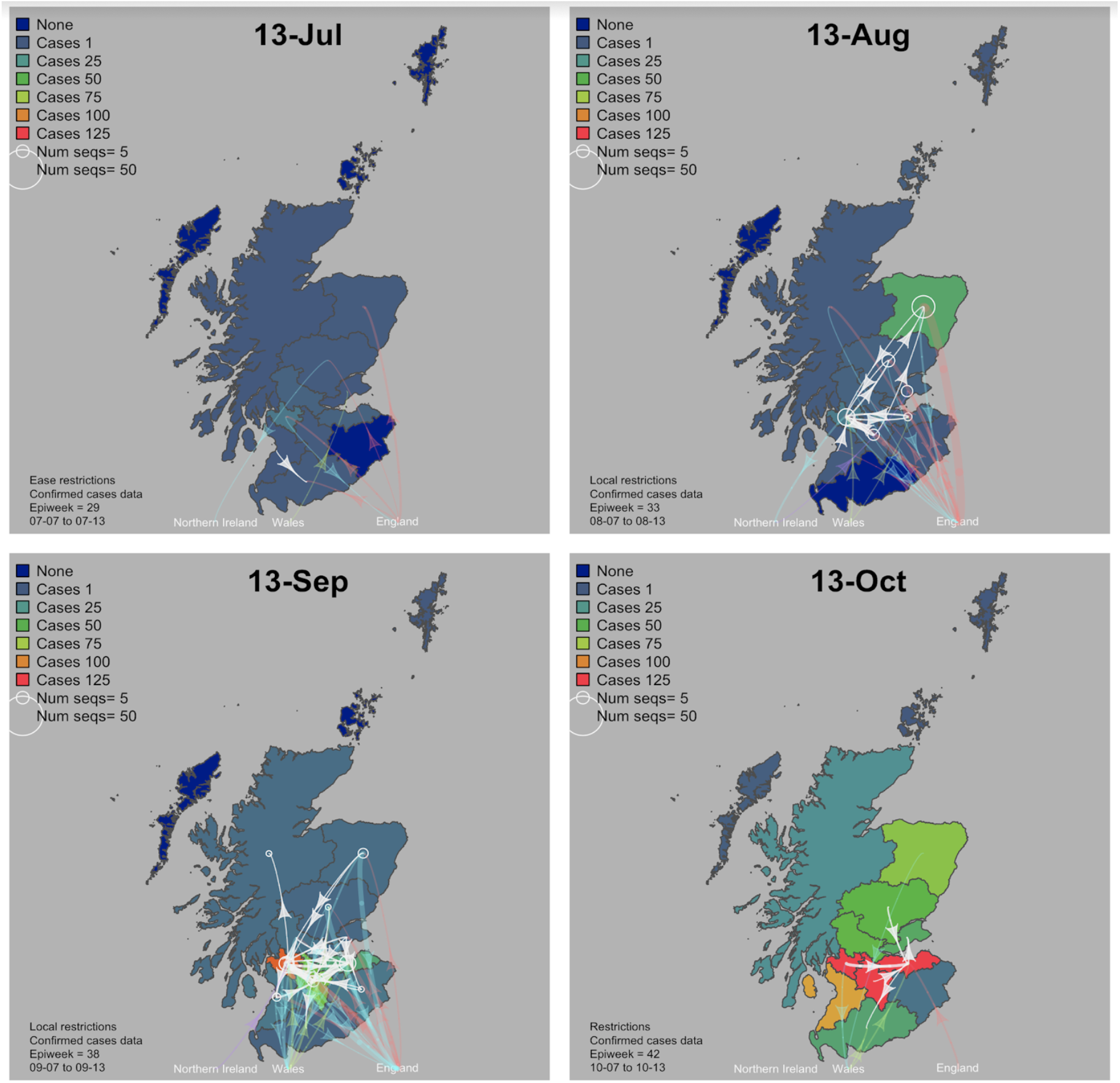
Maps showing the import and export relationships between the Health Boards within Scotland at different timepoints from July to October. A time lapse video is available at http://phylodynamics.lycett.roslin.ed.ac.uk:3838/RiseFallScotCOVID/ Results for Orkney, Shetland and the Western Isles are not included as sequence numbers are too low for these regions.

In order to estimate both when, and from where the UK lineages were imported into Scotland, time-scaled trees from the 21st Oct 2020 data set were used. As before, each UK lineage which had sequences in Scotland was ‘extended’ to include ancestral nodes and other sequences subtended from those nodes from the main global tree (see Methods). These extended lineages were time-scaled, and a discrete traits model was used to infer which region infected which. Here the simplest equal rates discrete trait model was used (most similar to parsimony). As previously noted, estimates can be biased by the number of sequences in each category, the accuracy of the sub trees, and additionally by the time-scaling mechanism. Although the sequence numbers only represent a few percent of the true number of cases (including untested individuals) adding additional identical sequences sampled in the same epi-week into a lineage will not change the estimate of the origin time of the lineage by very much, since this estimate is governed by the maximum diversity of the sequences.

Results for the ancestral origin of UK lineages that exist in Scotland (Supplementary figure 6 and Supplementary table 2) show that by July imports were increasing after the reduction in May and June, and there was an apparent peak of imports in August (although the estimation method is sensitive to having sufficient data subsequent to August to make this inference). We estimate that during the period 17th July to 30th August, there were 46 import events from elsewhere into Scotland, which were composed of 28 (61% of events) from England, 13 (28% of events) from Europe (not including UK), and four from Asia (9%). After August 30th, we estimate only nine import events from elsewhere into Scotland with eight of them having been detected in England and one from Europe. Considering that the number of sequences from England is by far the highest of any country or continent region, it is not surprising that there are so many apparent imports from England, but it is noteworthy that there are apparent detectable imports into Scotland from Europe and Asia in August. Furthermore, epidemiological investigations indicate that 11 of the 13 new lineages from epi-week 29 to 39 analysed contained sequences from individuals with recorded travel history mostly from Europe and a few from England. These independent epidemiological results largely agree with the results from the phylogenetic analyses that several lineages in Scotland were seeded by European imports and some by English imports over the summer period.

### The second wave -- relative contribution of imports to lineages in September

An estimate of the relative contribution of the imported lineages to the subsequent circulation in Scotland was made by considering which UK lineages were imported in the time periods 17th July to 30th August (summer) and 30th August onwards (autumn), and the number of sequences within those lineages in Scotland versus the total number of sequences in Scotland counting from 17th July onwards for summer imports, or 30th August onwards for autumn imports. Supplementary table 3 shows the summary per lineage origin location, and Supplementary tables 4 and 5 show the details of the top summer and autumn imported lineages.

There were 2882 sequences collected from Scotland from 17th July onwards, and of the 46 lineages which had an inferred origin outside Scotland timed between 17th July and 30th August, these lineages resulted in 763 sequences in Scotland, so overall it is estimated that imported lineages over the summer contributed to 26% of sequences from the summer onwards. Interestingly, by this measure it was imports from Europe (16%) rather than England (8%) that contributed more to the subsequent number of Scottish sequences over the summer and onwards. While, in the autumn, there were 1686 sequences collected from Scotland from 30th August onwards, and of the nine lineages which had an inferred origin outside Scotland timed as 30th August and onwards, these lineages resulted in 18 sequences in Scotland. So overall it is estimated that imported lineages over the autumn only contributed to 1% of sequences from the autumn onwards. Of this 1% the majority of the sequences in these lineages had an English origin.

## Conclusions

COVID-19 in Scotland was seeded by at least 300 introductions of SARS-CoV-2 across the country with the majority of imports being from our nearest neighbours England and mainland Europe. Scotland in turn exported the virus to multiple geographic locations, most notably England, North America and mainland Europe. Within Scotland itself the main metropolitan areas were greater contributors to the seeding of SARS-CoV-2 into other geographical locations. It is clear that due to the extent the virus was established in Scotland by April, without the lockdown infections would likely have continued to spread and rise uncontrolled. Notwithstanding the unintended consequences of the lockdown on individuals lives, other medical conditions, mental health, education and the economy, it can be concluded that this unprecedented public health intervention, not only reduced case numbers as is well documented, but led to reductions in the spread of the virus and to the extinguishing of multiple fast growing virus lineages. However, it is clear from the genomics data that it is imperative to control SARS-CoV-2 re-emergence appropriately once viral prevalence has been reduced to low levels. Prevention measures to minimise re-introductions into the UK and the virus being moved from regions of high to low prevalence are important. SARS-CoV-2 is established in the human population and until a vaccine or effective drugs are available, non-pharmaceutical interventions are all we have to control the virus, and therefore it is paramount that the lessons learned about their effectiveness are heeded for future public health benefit.

## Methods

### Data acquisition

SARS-CoV-2 positive samples were obtained by the Royal Infirmary of Edinburgh and the MRC-University of Glasgow Centre for Virus Research (CVR) from the eleven Health Boards of Scotland. Viral genome sequences were obtained (see below), combined with others from across the UK and with genomes from other countries also deposited in GISAID (Supplementary table 1). All known high quality genomes in GISAID were initially processed. As of 2020-08-18 these consist of 72401 genomes in total, of which 35744 were from the UK (approximately 50% of the global sample) and 5137 were from Scotland (7% of the global sample, and 14% of the whole UK sample; and 26.5% of the total positive tests in Scotland). The global data set contained sequences sampled from 2019-12-24 to 2020-08-10, of which Scottish sequences were from 2020-02-28 to 2020-08-05, the later a few weeks after the time lockdown restrictions started to be relaxed (taken to be 2020-07-10). To study the second wave SARS-CoV-2 genome data available until the 2020-10-21 was analysed.

### Samples

Samples were obtained from all Scottish Health Boards following ethical approval from the relevant national biorepository authorities (16/WS/0207NHS). The Royal Infirmary of Edinburgh and West of Scotland Specialist Virology Centre, NHS GGC conducted diagnostic real-time RT-PCR to detect SARS-CoV-2 positive samples, following nucleic acid extraction. Residual nucleic acid from samples were sequenced at the Royal Infirmary of Edinburgh and the MRC-University of Glasgow Centre for Virus Research (CVR) for whole genome next generation sequencing.

### Sequencing, ONT MinION/GridION

Following extraction, libraries were prepared utilising protocols developed by the ARTIC network (v1 and v2) and primer versions v1-v3. The sequencing protocol and multiplex primers are described in detail at https://artic.network/ncov-2019. Library pools were loaded onto R9.4.1 flow cells and sequenced in MinION or GridION devices using MinKNOW version 19.12.5 and 19.12.6, respectively. The ARTIC nCoV-2019 environment utilising RAMPART v1.0.5 and v1.1.0 was used to visualise read mapping in real time. MinION generated raw FAST5 files were subsequently base called using Guppy version 3.4.5 in high accuracy mode and using a minimum quality score of 7. GridION generated fastq files were also produced utilising live high accuracy base calling. Following the ARTIC-nCov-2019 bioinformatics protocol, reads were initially size filtered, then demultiplexed and trimmed with Porechop (18), and mapped against reference strain Wuhan-Hu-1 (GenBank accession number MN908947.3), followed by clipping of primer regions. Variants were called using Nanopolish 0.11.3 (19) and accepted if they had a log-likelihood score of greater than 200, and read coverage of at least 20.

### Sequencing, Illumina MiSeq

Samples were processed as described for the ONT workflow, until the amplicon generation stage. The resulting DNA fragments were cleaned using AMPURE beads (Beckman Coulter) and used to prepare Illumina sequencing libraries with a DNA KAPA library preparation kit (Roche) following manufacturer’s instructions. Indexing was carried out with NEBNext multiplex oligos (NEB), using 7 cycles of PCR. Libraries were pooled in equimolar amounts and loaded on a MiSeqV2 cartridge (500 cycles). The PrimalAlign pipeline (https://github.com/rjorton/PrimalAlign) was used to construct the consensus. A read coverage of at least 10 was used for the consensus. Some of the samples were sequenced at the Wellcome Sanger Institute, following the protocol described in detail at protocols.io (https://www.protocols.io/view/covid-19-artic-v3-illumina-library-constructionan-bibtkann; https://www.protocols.io/view/covid-19-artic-v3-illumina-library-construction-anbnidmca6. Briefly, RNA was reversed transcribed using LunaScript RT (NEB) followed by amplicon generation using NEB Q5® Hot Start High-Fidelity (NEB) and the ARTIC network primers. Following clean up, libraries were prepared using the NEBNext Ultra II DNA Library Prep Kit for Illumina (NEB), including end prep, ligation of a TruSeq adapter to the DNA fragments, followed by PCR amplification and indexing with KAPA 2x Kapa HiFi Hotstart (Roche), and KAPA unique dual indices (Roche). Following quantification and pooling, libraries were loaded on a Illumina NovaSeq SP flow cell, using the XP workflow, with 384 samples per lane.

### Sequence data

Only consensus sequences >90% coverage were included in our analysis. All consensus genomes are available from the GISAID database (https://www.gisaid.org), the COG-UK consortium website (https://www.cogconsortium.uk/data/) and BAM files from the European Nucleotide Archive’s Sequence Read Archive service, BioProject PRJEB37886 (https://www.ebi.ac.uk/ena/data/view/PRJEB37886).

### Phylogenetic analysis

Global sequences were obtained from the GISAID Initiative https://www.gisaid.org. All UK and global sequences were processed using the COG-UK grapevine pipeline https://github.com/COG-UK/grapevine. Briefly, the sequences are aligned to the reference sequence Wuhan-Hu_1 (MN908947.3) using minimap2 (Li, 2018) and a maximum-likelihood phylogenetic tree is constructed using Fasttree http://www.microbesonline.org/fasttree/. PANGOLIN, https://pangolin.cog-uk.io (Rambaut et al. 2020), was used for assigning phylogenetic lineages. UK lineages are defined here: https://github.com/COG-UK/grapevine/blob/master/docs/lineages.md. Note, the UK lineage numbering is dynamic so can change with data updates. TreeTime (Sagulenko, Puller, and Neher 2018) was used to estimate a time calibrated phylogeny for all Scottish sequences up until 2020-08-18 using a clock rate of 0.001 allowing for up to 10 iterations in the inference cycle. Faith’s phylogenetic diversity (Faith 1992) was used to estimate phylogenetic diversity by summing the length of the branches for each NHS Health Board for each epi-week.

### Phylodynamic analysis

For each UK lineage present in Scotland, the most-recent-common-ancestor (MRCA) internal node in the tree corresponding to the origin of the Scottish sequences was found. Next an ‘extended’ lineage was formed by going two steps backward in the tree and then re-extracting the original lineage plus a subsample of the other sequences subtended by that ancestral node, resulting in the original clade of Scottish sequences embedded in surrounding sequences from elsewhere in the UK and globally. Polytomies in the extended lineage were resolved by including very small branch lengths (genetic distances of 1×10-12). An approximate time scale for this extended lineage tree was estimated using R package Treedater (Volz and Frost 2017). The estimation should be better than just estimating on the original lineage because the time-depth of the samples is greater. The clade in the extended lineage containing sequences only from Scotland was subsequently re-extracted from the time-scaled tree, and the skygrowth package in R was used to estimate the effective population size over time, and the growth rate of the lineage within Scotland (data shown in R-shiny). Recent travel history for a sample of wave two infections were obtained from local public health databases.

## Supporting information

COG-UK Authorship list

GISAID acknowledgment file

## Data Availability

We thank the Scottish NHS virology laboratories who provided samples for sequencing, the authors who have shared genome data on GISAID, https://www.gisaid.org. Our genome sequence acknowledgments can be found in the Supplementary. Online results of the analyses are displayed in http://sars2.cvr.gla.ac.uk/RiseFallScotCOVID/ (data to 2020-08-18) and http://phylodynamics.lycett.roslin.ed.ac.uk:3838/RiseFallScotCOVID/ (data to 2020-10-21)
This study focuses on public health/surveillance questions in Scotland that make use of sequence data and other metadata that was collected under the activities of the COG-UK consortium (https://www.cogconsortium.uk/). COG-UK data is released and is publicly available via the ENA, GISAID and the COG-UK website.

http://phylodynamics.lycett.roslin.ed.ac.uk:3838/RiseFallScotCOVID/

## Acknowledgements

We thank the Scottish NHS virology laboratories who provided samples for sequencing, the authors who have shared genome data on GISAID, https://www.gisaid.org. Our genome sequence acknowledgments can be found in Supplementary table 6. We thank Scott Arkison for HPC upgrades and systems administration. Funding: MRC (MC UU 1201412), Wellcome Trust Collaborator Award (206298/Z/17/Z – ARTIC Network), TCW Wellcome Trust Award 204802/Z/16/Z, Chief Scientist Office Project (COV/EDI/20/11), MRC EAVE II (MR/R008345/1), UKRI Industrial Strategy Challenge Fund BREATHE (MC_PC_19004), Scottish Government DG Health and Social Care, and Tommy’s Charity (1060508, SC03928). COG-UK is supported by funding from the Medical Research Council (MRC) part of UK Research & Innovation (UKRI), the National Institute of Health Research (NIHR) and Genome Research Limited, operating as the Wellcome Sanger Institute. CLIMB is funded by the Medical Research Council (MRC) through grant MR/L015080/1. SL additionally received support from Scottish Government Rural and Environment Science and Analytical Services Division as part of Centre of Expertise on Animal Disease Outbreaks (EPIC), and the BBSRC Institute Strategic Programme grant to Roslin Institute (BB/J004235/1).

