## Supplementary figures for "Epidemic waves of COVID-19 in Scotland: a genomic perspective on the impact of the introduction and relaxation of lockdown on SARS-CoV-2"

### Supplementary Material

Interactive versions of plots with data to 2020-08-18 (first wave) can be found at R-shiny RiseFallScotCOVID app: http://sars2.cvr.gla.ac.uk/RiseFallScotCOVID/

Interactive versions of plots with 2020-10-21 data (second wave) can be found at R-shiny RiseFallScotCOVID app: http://phylodynamics.lycett.roslin.ed.ac.uk:3838/RiseFallScotCOVID/

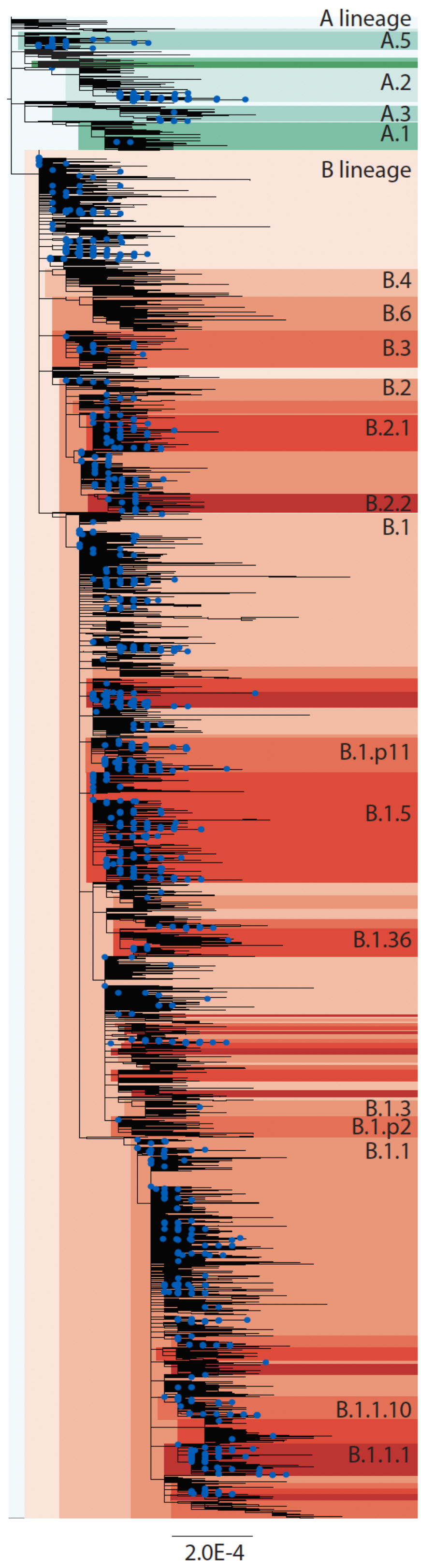


**Supplementary figure 1**. Phylogenetic trees showing the SARS-CoV-2 diversity circulating in Scotland August 18th 2020 comparison to the global lineages defined by (Rambaut et al. 2020). Scottish variants are denoted with blue circles.

A

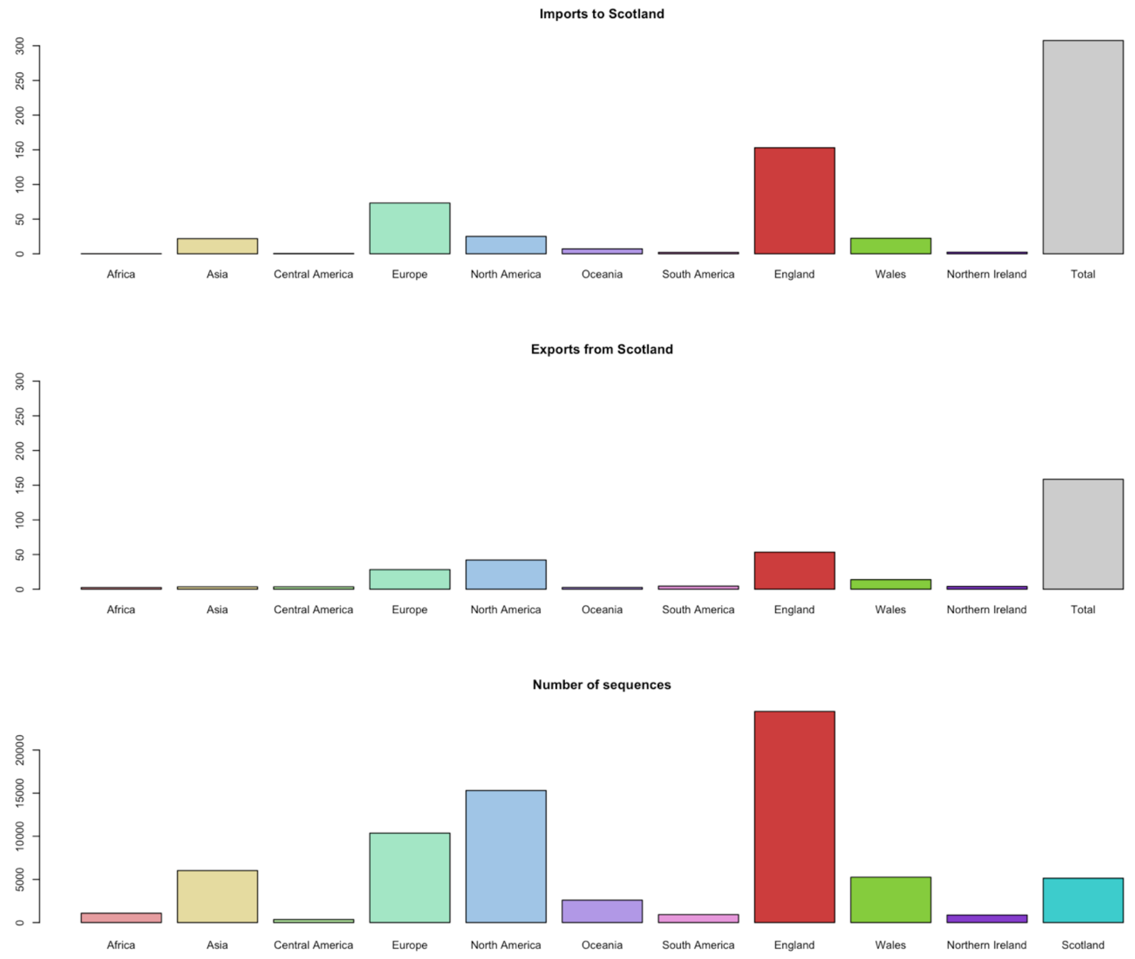


B

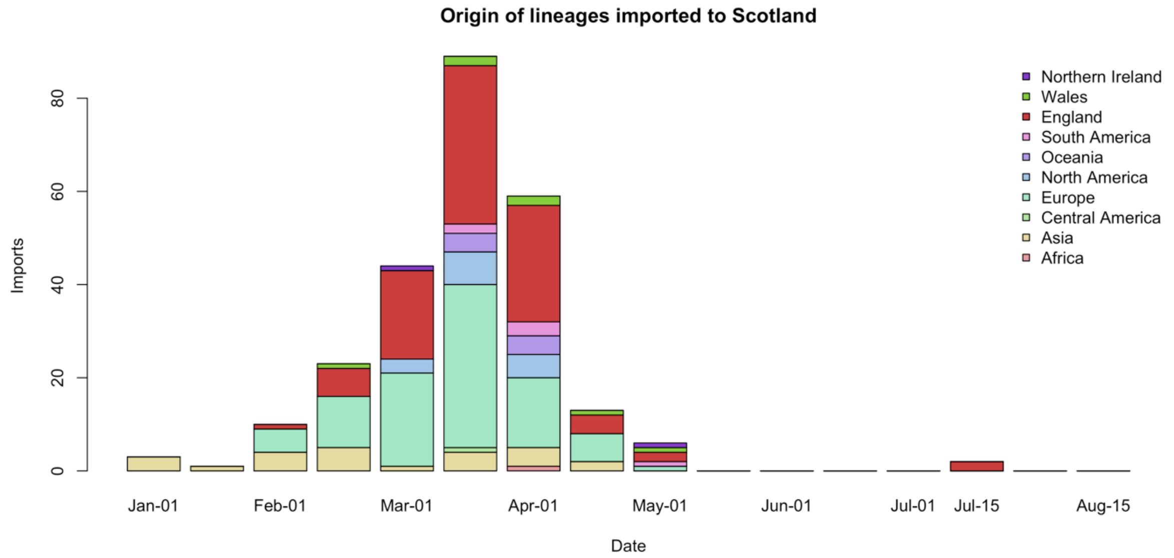


**Supplementary figure 2**. Estimate of the number of imports and exports of SARS-CoV-2 cases into Scotland using data collected up to 18th August 2020. A| by geographical region and B| lineage imports by time.

#### A - Ayrshire and Arran

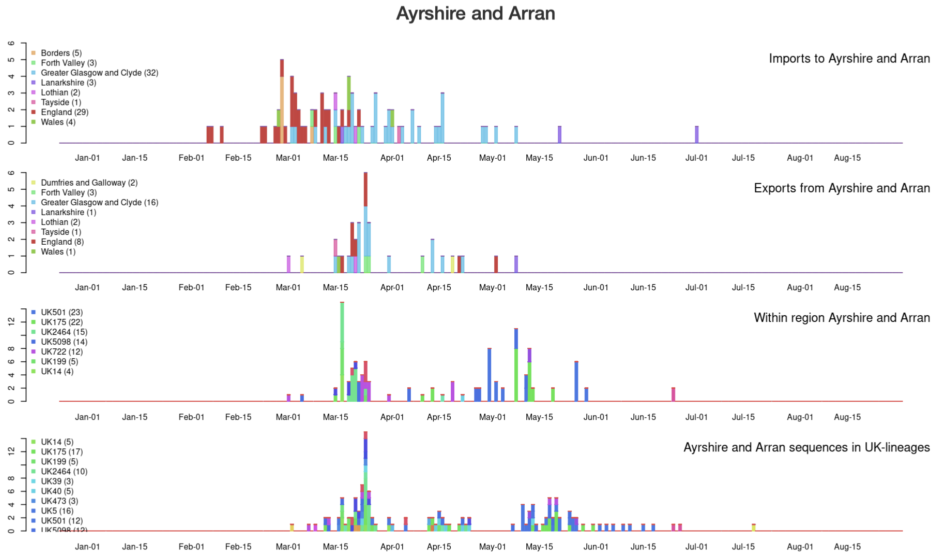


##### Summary for Health Board

| | Pre | Mid | Post |
|---|---|---|---|
| Imports | 53.00 | 26.00 | 0.00 |
| Exports | 15.00 | 19.00 | 0.00 |
| Within | 41.00 | 87.00 | 0.00 |
| Samples | 27.00 | 104.00 | 0.00 |

#### B - Borders

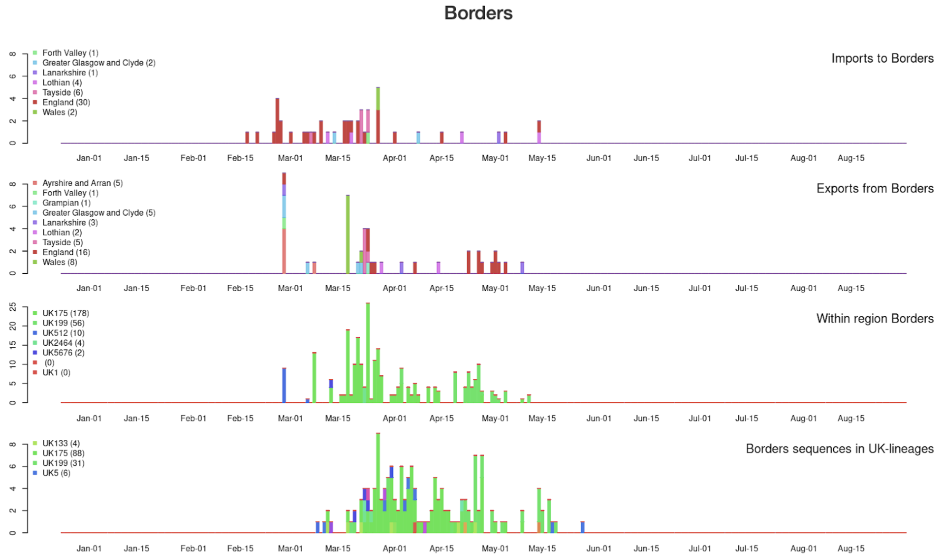


##### Summary for Health Board

| | Pre | Mid | Post |
|---|---|---|---|
| Imports | 30.00 | 16.00 | 0.00 |
| Exports | 25.00 | 21.00 | 0.00 |
| Within | 95.00 | 155.00 | 0.00 |
| Samples | 13.00 | 139.00 | 0.00 |

#### C - Dumfries and Galloway

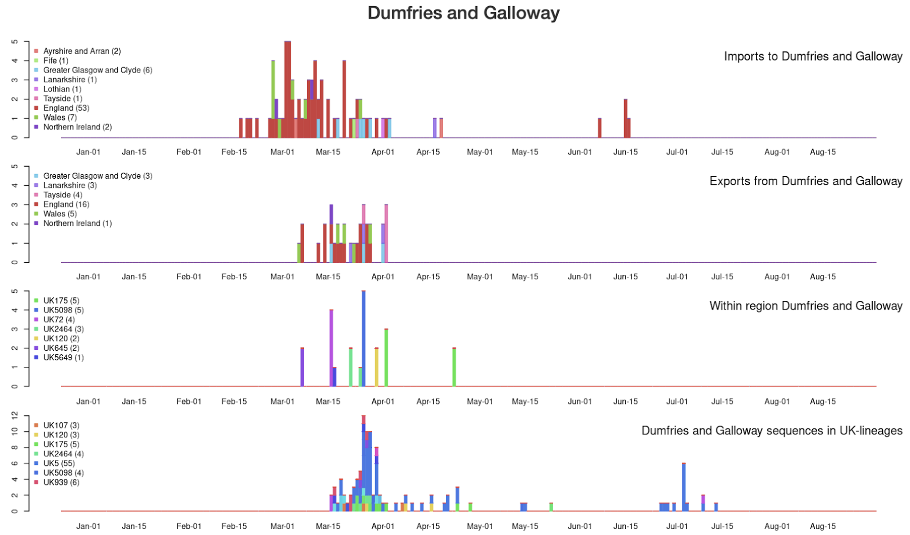


##### Summary for Health Board

| | Pre | Mid | Post |
|---|---|---|---|
| Imports | 56.00 | 18.00 | 0.00 |
| Exports | 16.00 | 16.00 | 0.00 |
| Within | 44.00 | 40.00 | 0.00 |
| Samples | 15.00 | 90.00 | 0.00 |

### D - Fife

#### Fife

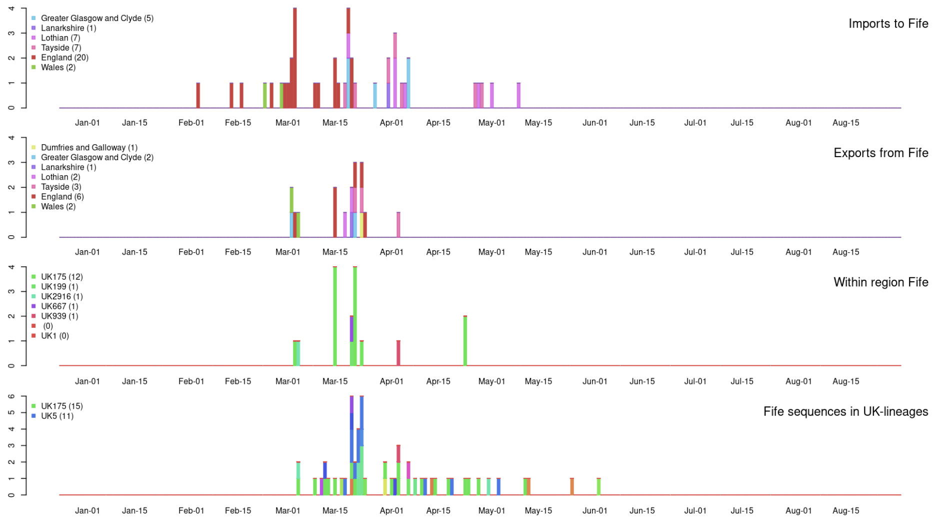


#### Summary for Health Board

| | Pre | Mid | Post |
|---|---|---|---|
| Imports | 27.00 | 15.00 | 0.00 |
| Exports | 14.00 | 3.00 | 0.00 |
| Within | 30.00 | 5.00 | 0.00 |
| Samples | 22.00 | 32.00 | 0.00 |

### E - Forth Valley

#### Forth Valley

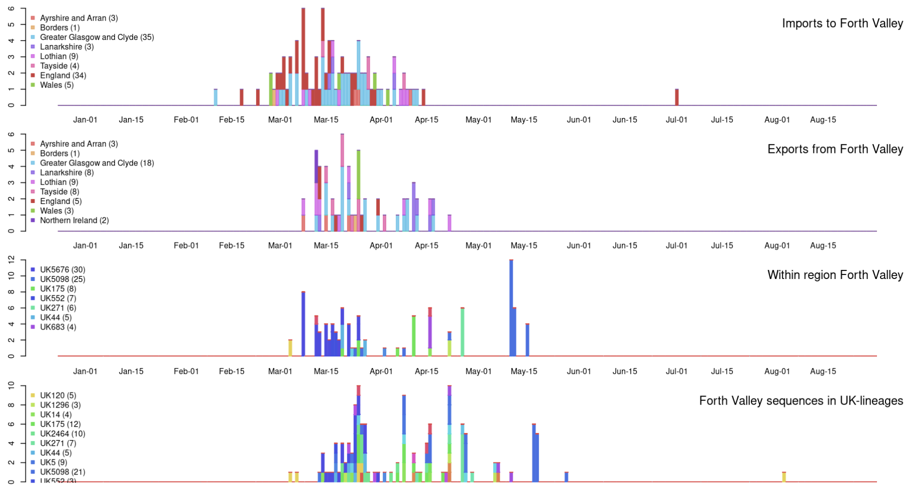


#### Summary for Health Board

| | Pre | Mid | Post |
|---|---|---|---|
| Imports | 62.00 | 32.00 | 0.00 |
| Exports | 30.00 | 27.00 | 0.00 |
| Within | 48.00 | 65.00 | 0.00 |
| Samples | 24.00 | 105.00 | 1.00 |

### F - Grampian

#### Grampian

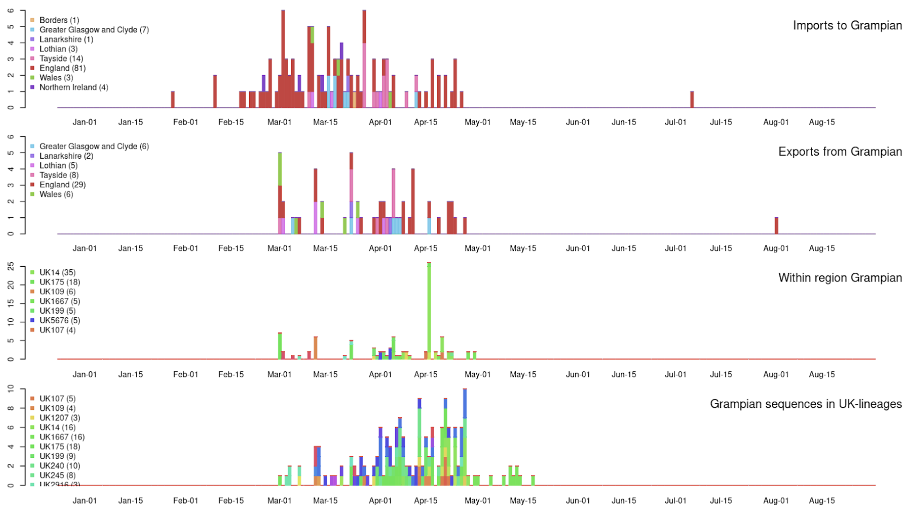


#### Summary for Health Board

| | Pre | Mid | Post |
|---|---|---|---|
| Imports | 72.00 | 42.00 | 0.00 |
| Exports | 20.00 | 35.00 | 1.00 |
| Within | 30.00 | 101.00 | 91.00 |
| Samples | 19.00 | 145.00 | 0.00 |

### G - Greater Glasgow and Clyde

#### Greater Glasgow and Clyde

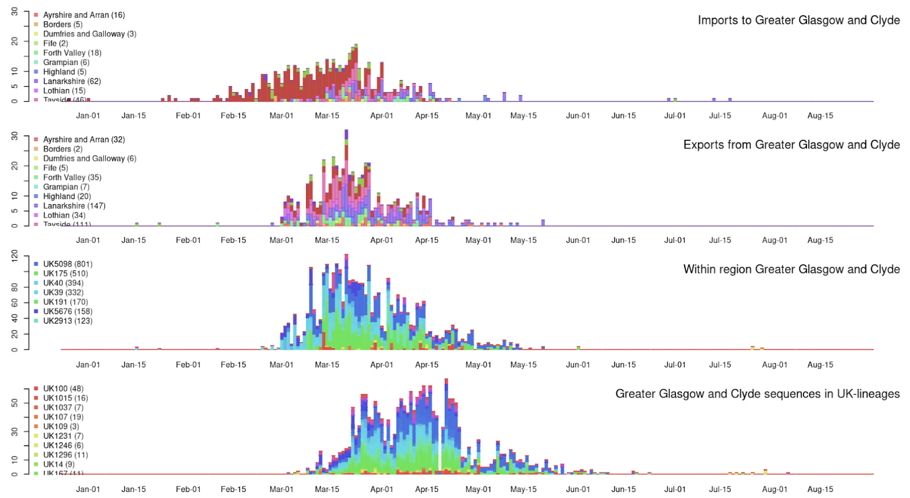


##### Summary for Health Board

| | Pre | Mid | Post |
|---|---|---|---|
| Imports | 339.00 | 167.00 | 0.00 |
| Exports | 287.00 | 250.00 | 0.00 |
| Within | 1468.00 | 1854.00 | 6.00 |
| Samples | 143.00 | 1821.00 | 7.00 |

### H - Highland

#### Highland

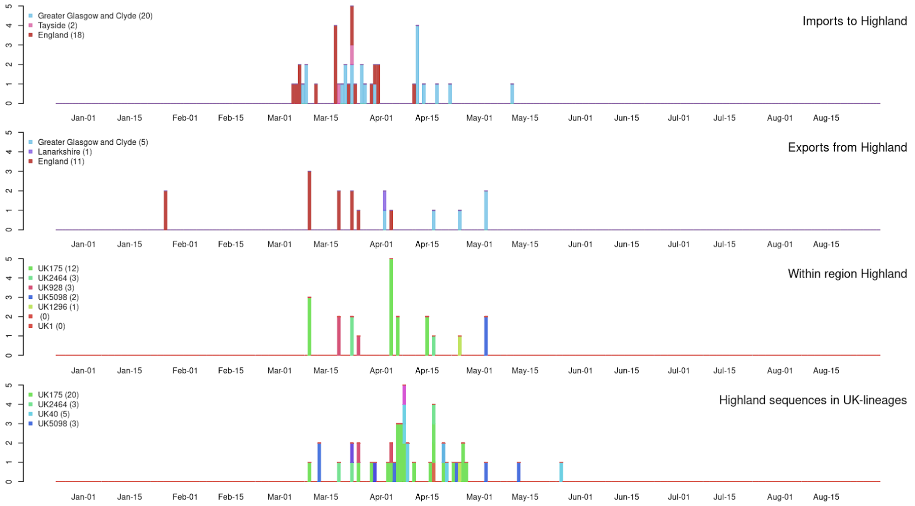


##### Summary for Health Board

| | Pre | Mid | Post |
|---|---|---|---|
| Imports | 20.00 | 20.00 | 0.00 |
| Exports | 7.00 | 10.00 | 0.00 |
| Within | 7.00 | 14.00 | 0.00 |
| Samples | 4.00 | 41.00 | 0.00 |

### I - Lanarkshire

#### Lanarkshire

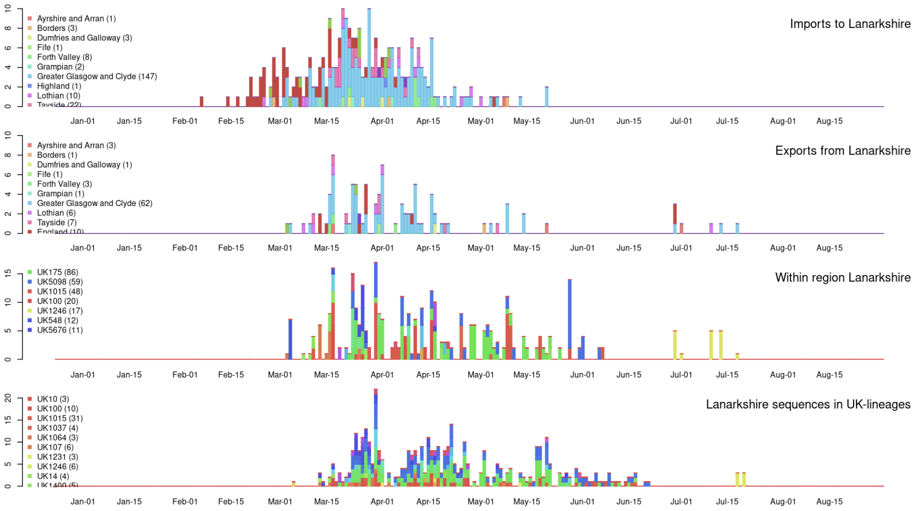


##### Summary for Health Board

| | Pre | Mid | Post |
|---|---|---|---|
| Imports | 125.00 | 142.00 | 0.00 |
| Exports | 28.00 | 70.00 | 0.00 |
| Within | 64.00 | 296.00 | 0.00 |
| Samples | 22.00 | 381.00 | 3.00 |

#### J - Lothain

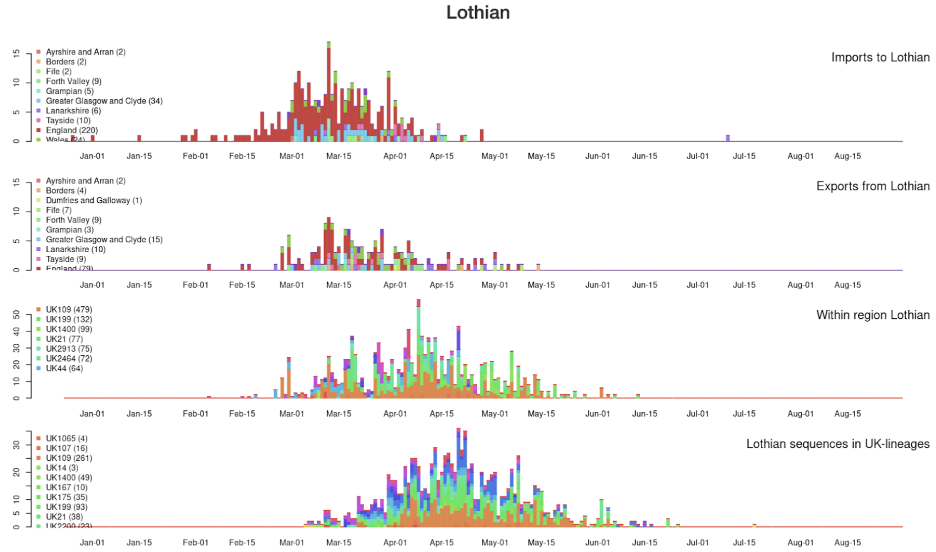


##### Summary for Health Board

| | Pre | Mid | Post |
|---|---|---|---|
| Imports | 238.00 | 80.00 | 0.00 |
| Exports | 95.00 | 74.00 | 0.00 |
| Within | 339.00 | 1182.00 | 0.00 |
| Samples | 63.00 | 986.00 | 0.00 |

#### K - Tayside

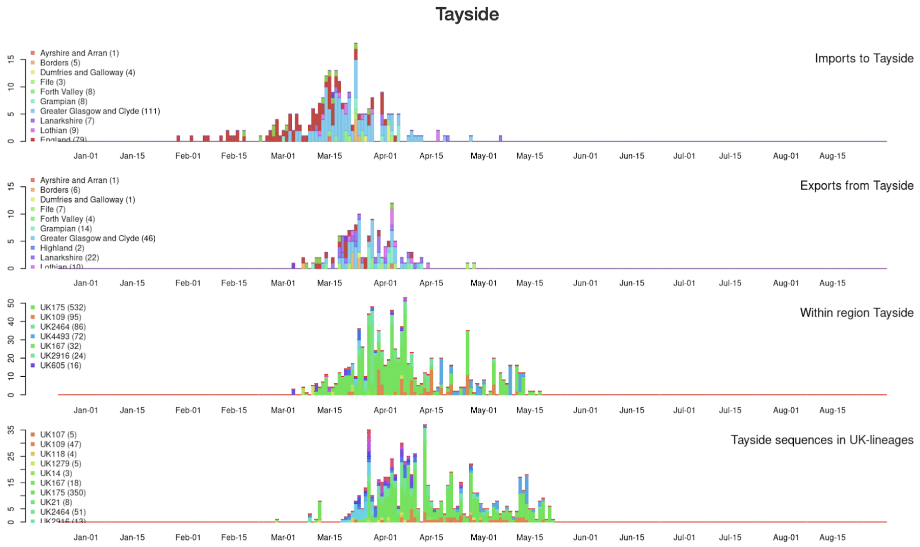


##### Summary for Health Board

| | Pre | Mid | Post |
|---|---|---|---|
| Imports | 173.00 | 75.00 | 0.00 |
| Exports | 53.00 | 84.00 | 0.00 |
| Within | 129.00 | 812.00 | 0.00 |
| Samples | 23.00 | 655.00 | 0.00 |

**Supplementary figure 3.** Timing of the imports and exports to Scottish Health Boards for data to 2020-08-18 (see Methods). Results for Orkney, Shetland and the Western Isles are not included as there are too few sequences. The four plots for each NHS Health Board are as follows: top plot -- imports coloured by first NHS Health Board lineage detected in (rainbow colours) or England (red), Wales (green), Northern Ireland (purple). Note that the England (red) is a major source for both NHS Greater Glasgow and Clyde and Lothian in March; second row plot -- exports (same colouring as imports); third row plot -- within region transmissions coloured according to UK lineage number (other rainbow colours); bottom plot -- number of sequences per NHS Health Board coloured by UK lineage number (same as third row). For the summary tables, pre is before 23 March 2020, mid is 23 March - 10 July 2020, and post is after 10 July 2020.
Interactive versions available at http://sars2.cvr.gla.ac.uk/RiseFallScotCOVID/.

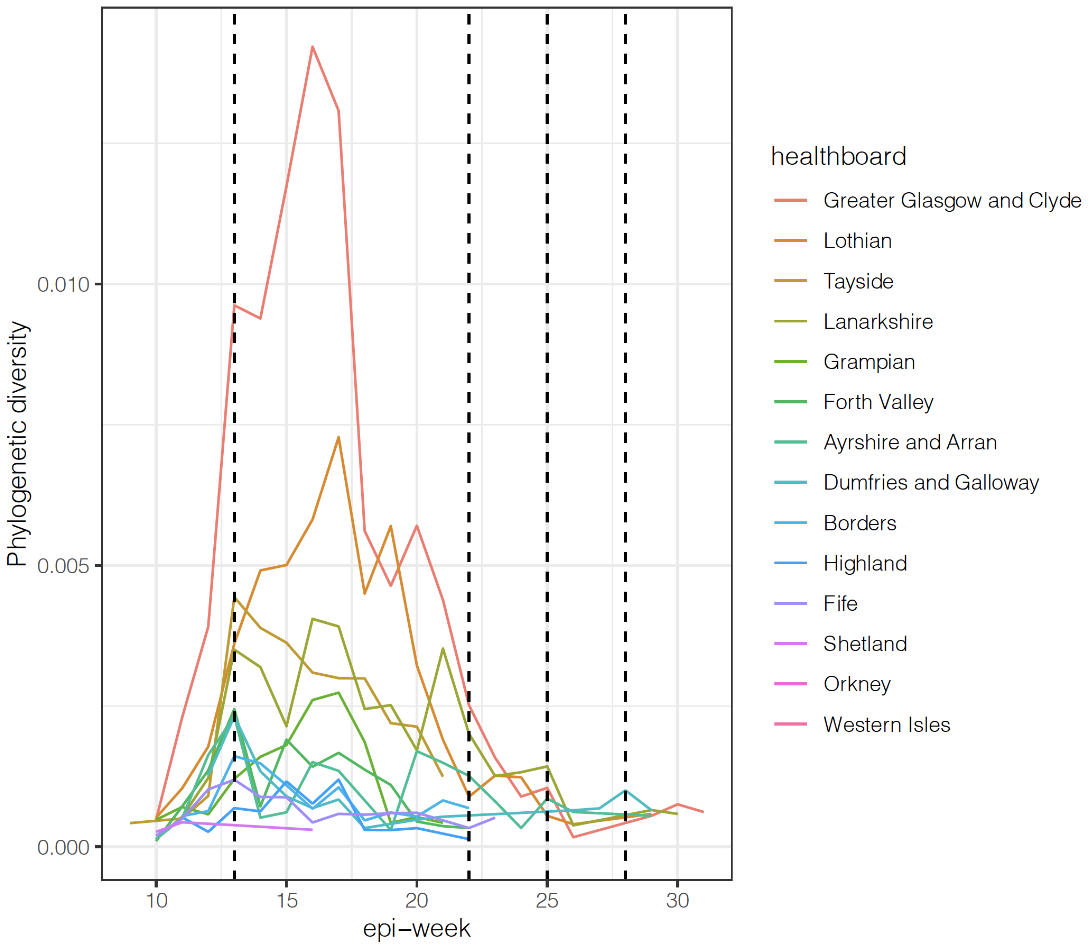


**Supplementary figure 4**. Phylogenetic diversity plotted against epi-week for each Scottish Health Board. The four vertical dashed lines represent the beginning of the lockdown (23rd March 2020) and its easing: movement to phase 1 (29th March 2020), phase 2 (19th June 2020) and phase 3 (10th July 2020).

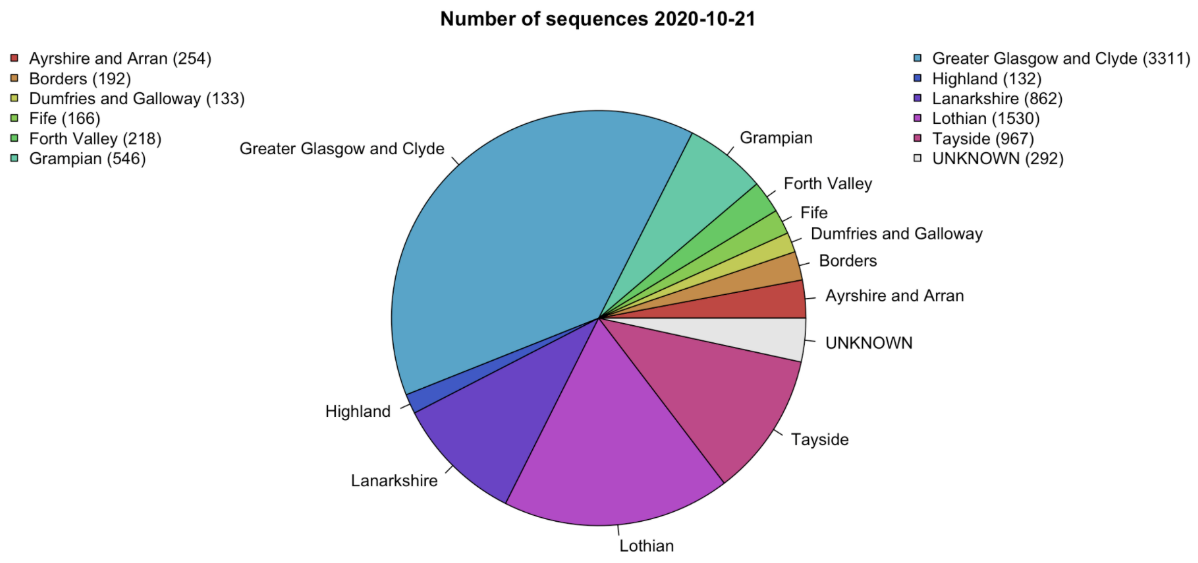


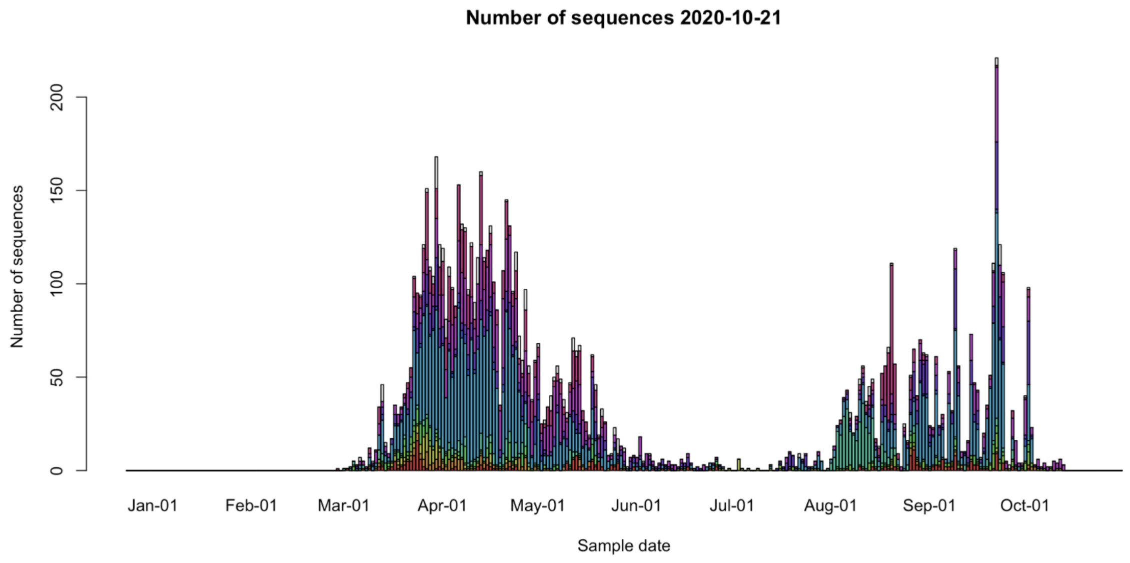


**Supplementary figure 5.** Sequence counts by Scottish Health Board (top) and by sampling time (bottom) to 21st October 2020.

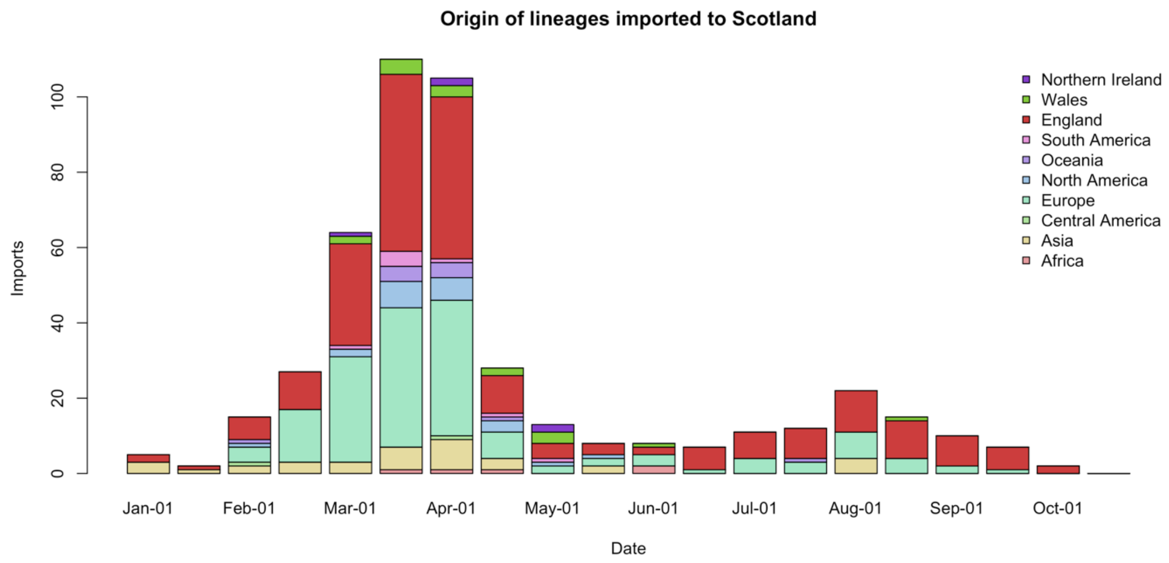


**Supplementary figure 6**. Estimate of the origin and timing of imports into Scotland using data to 21st October 2020.

#### A – Ayrshire and Arran

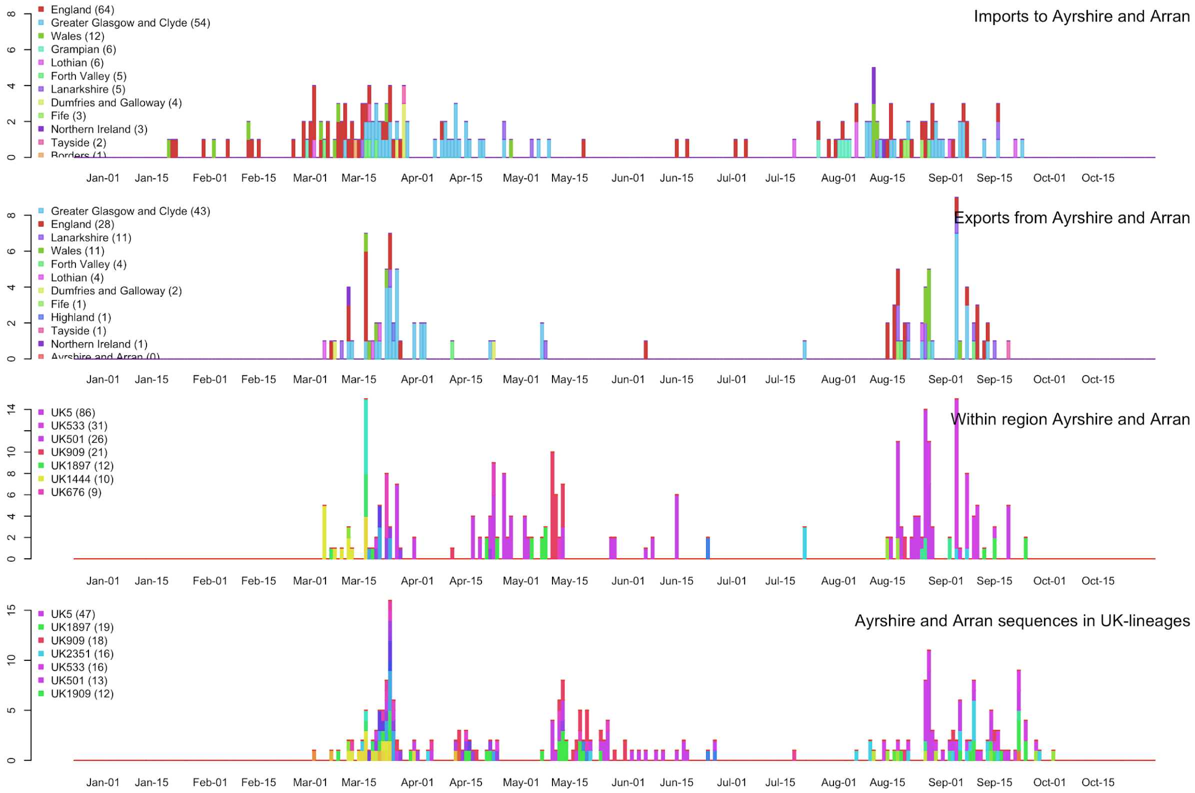


#### B - Borders

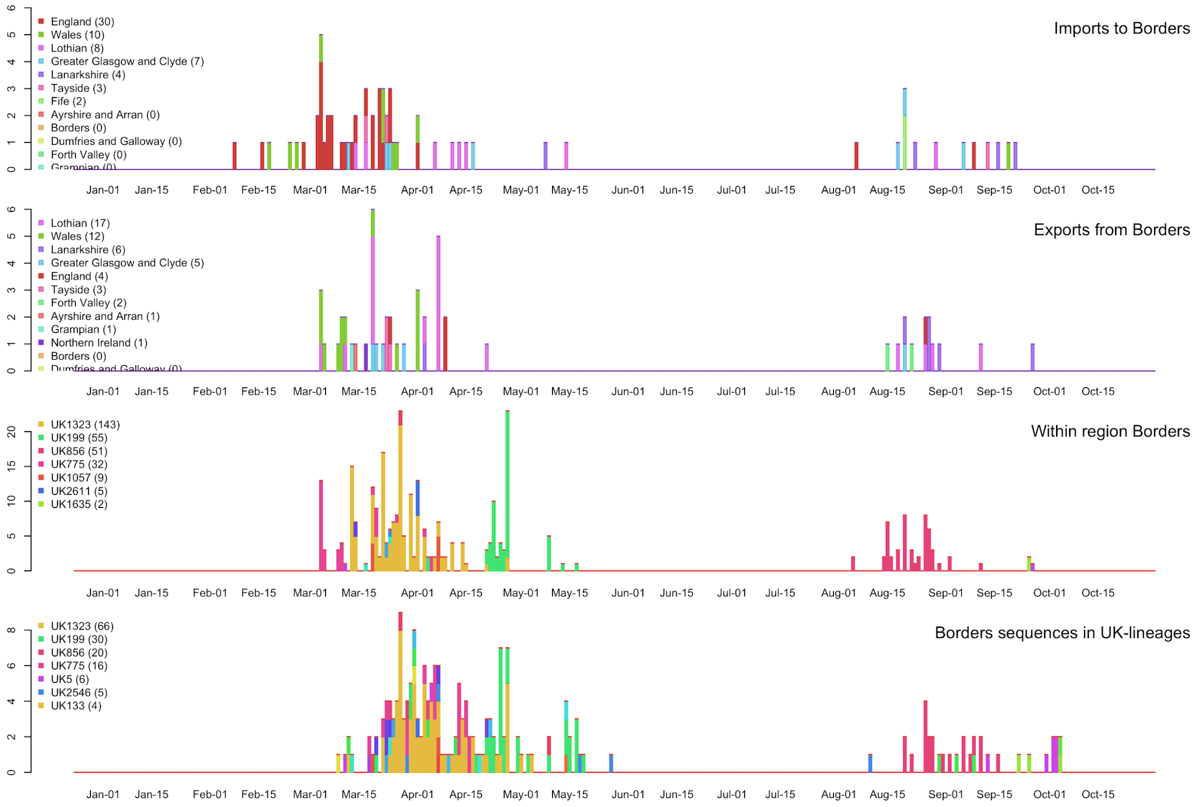

### C – Dumfries and Galloway

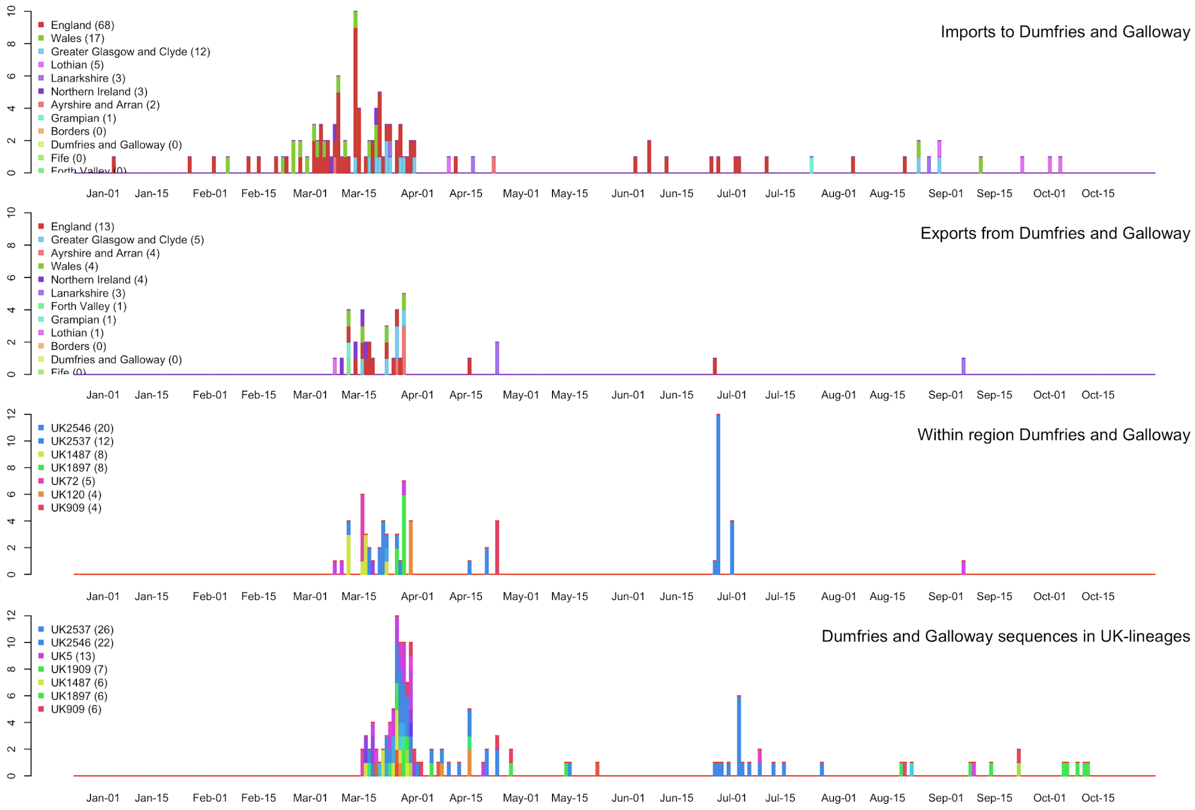


### D – Fife

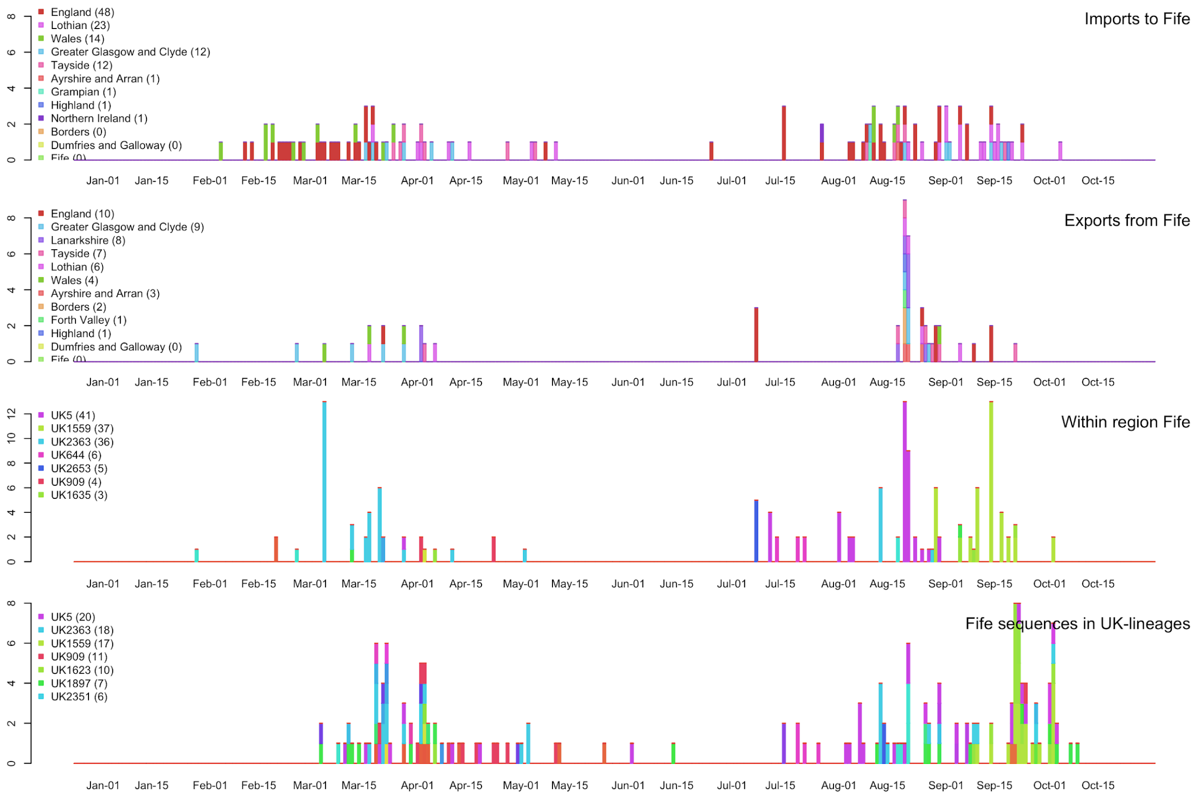

#### E – Forth Valley

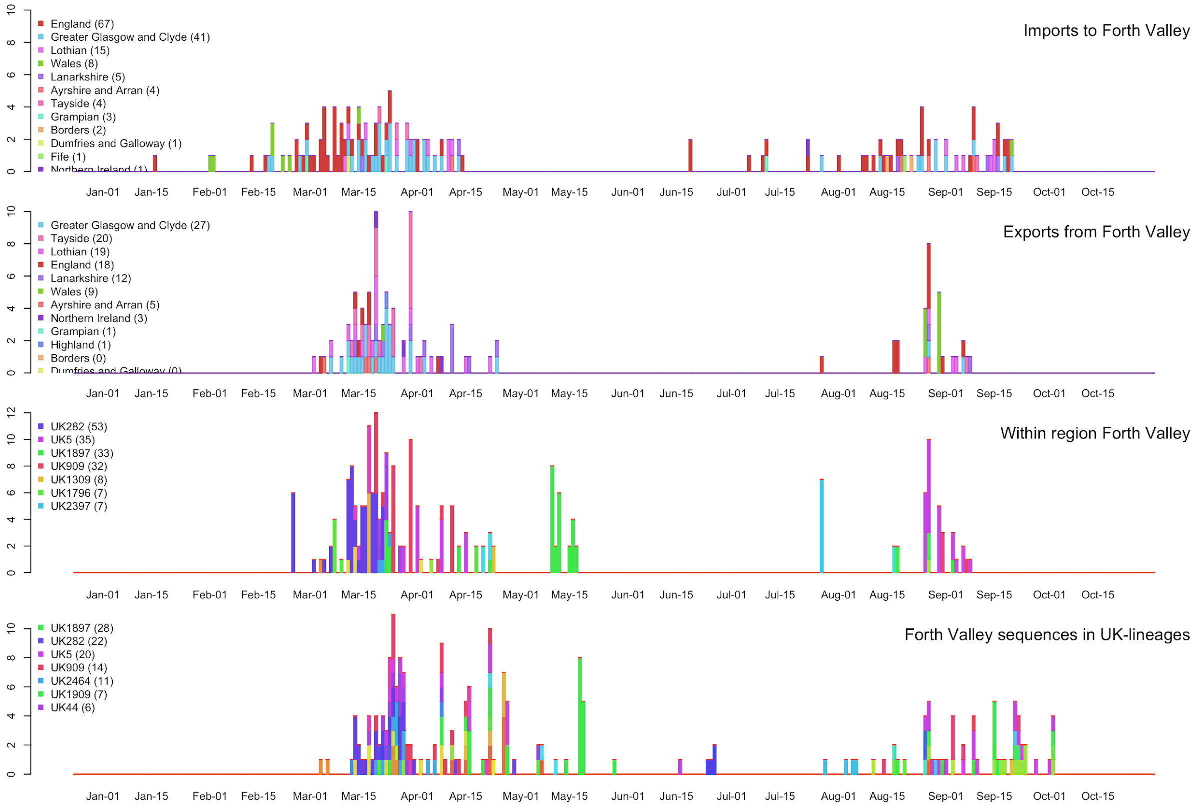


#### F – Grampian

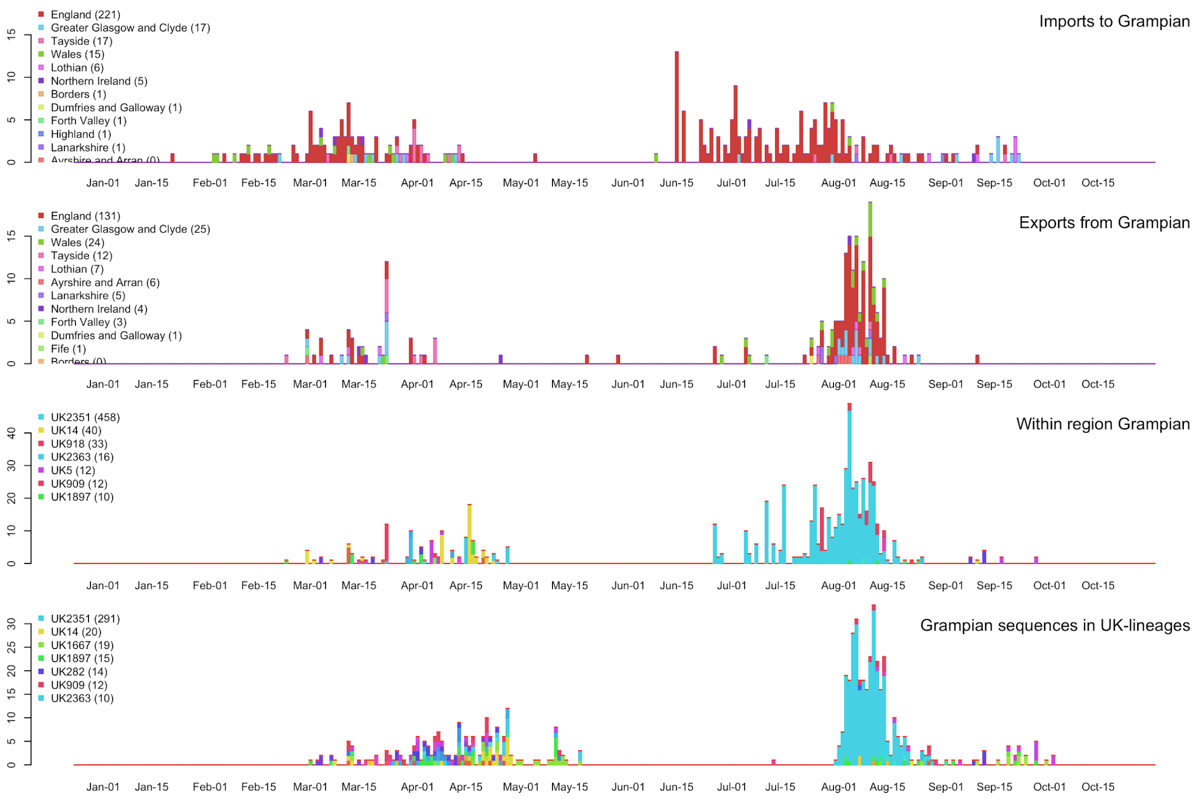

#### G – Greater Glasgow and Clyde

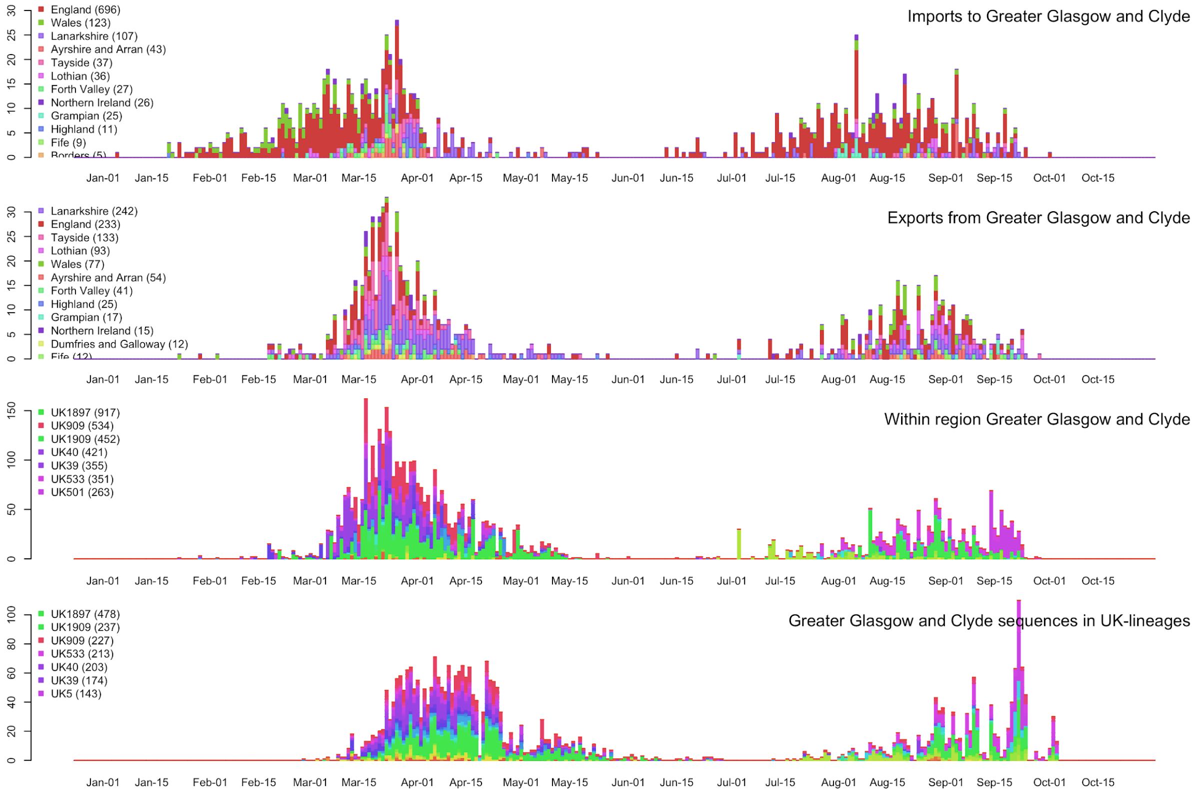


#### H - Highland

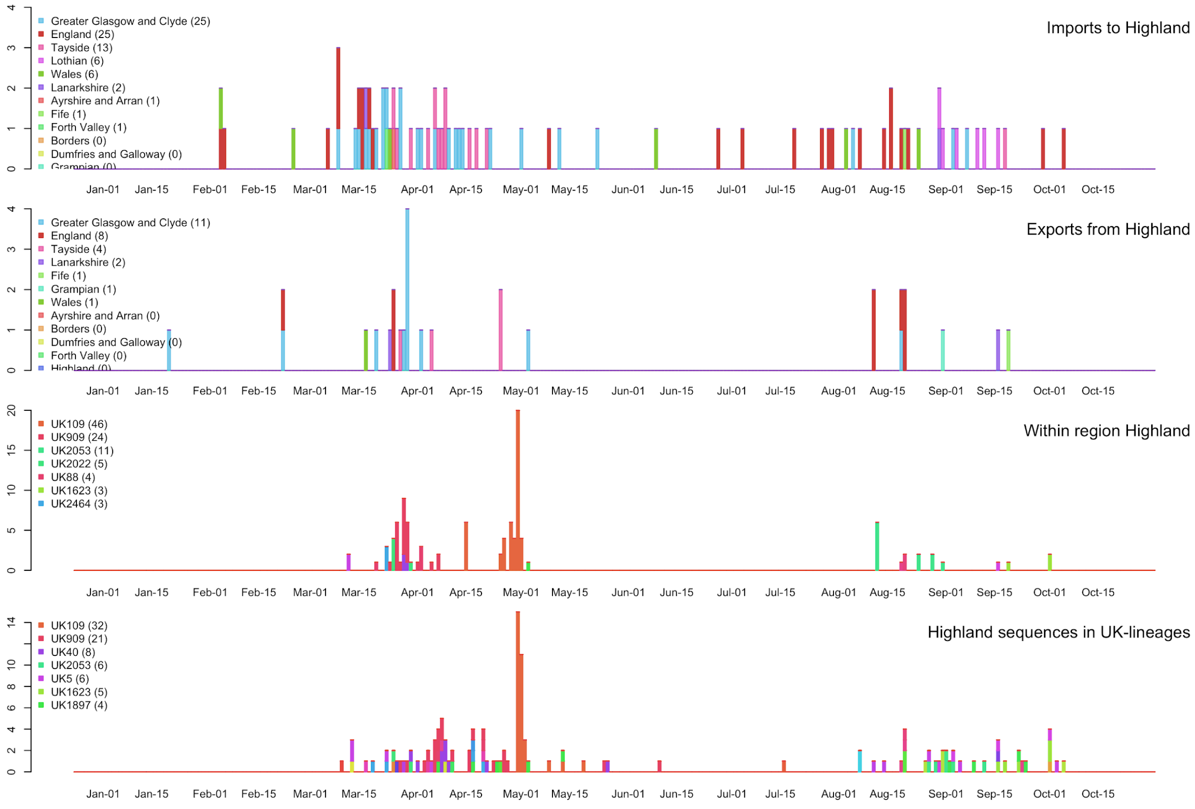

### I - Lanarkshire

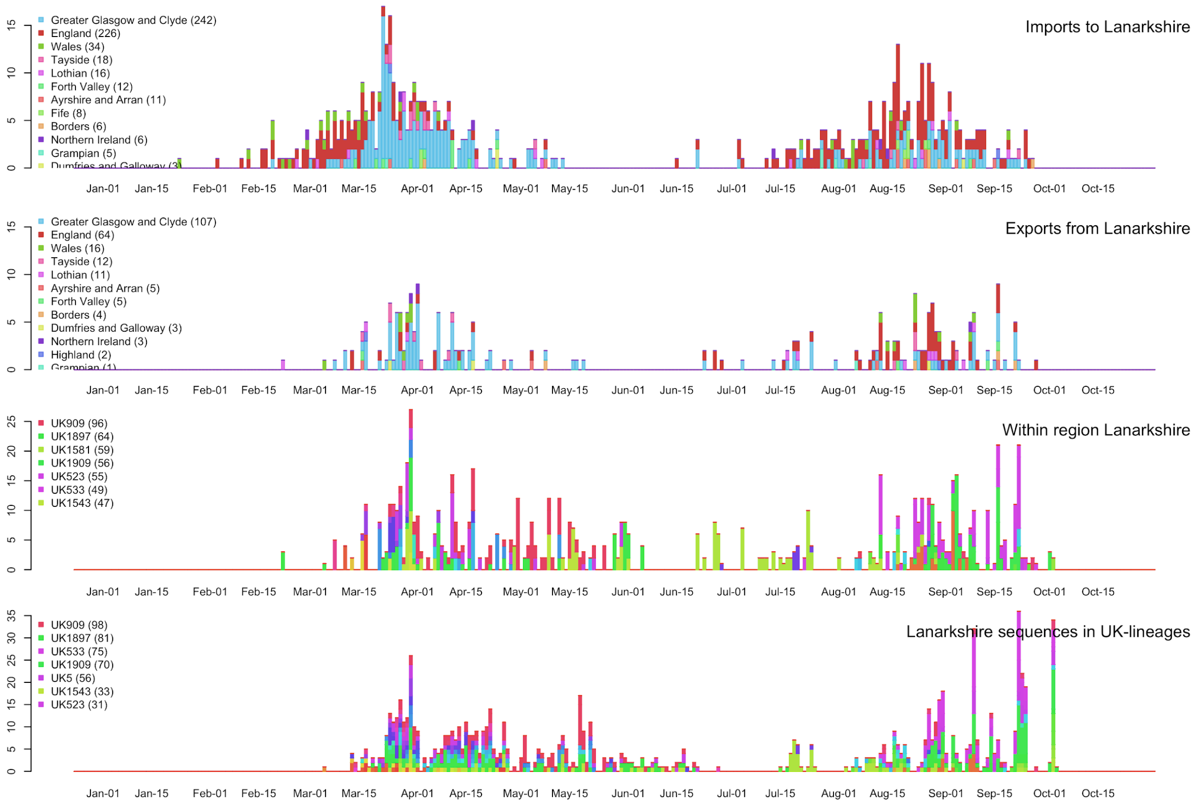


### J - Lothian

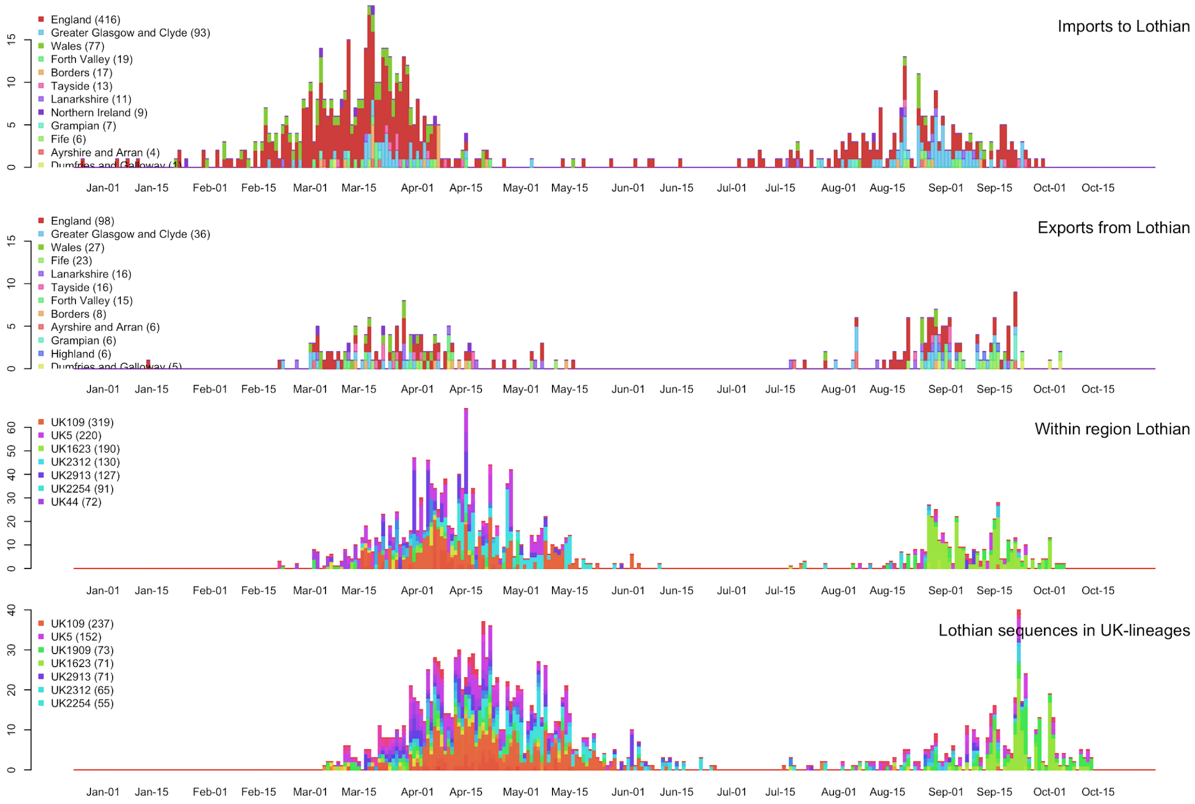

#### K – Tayside

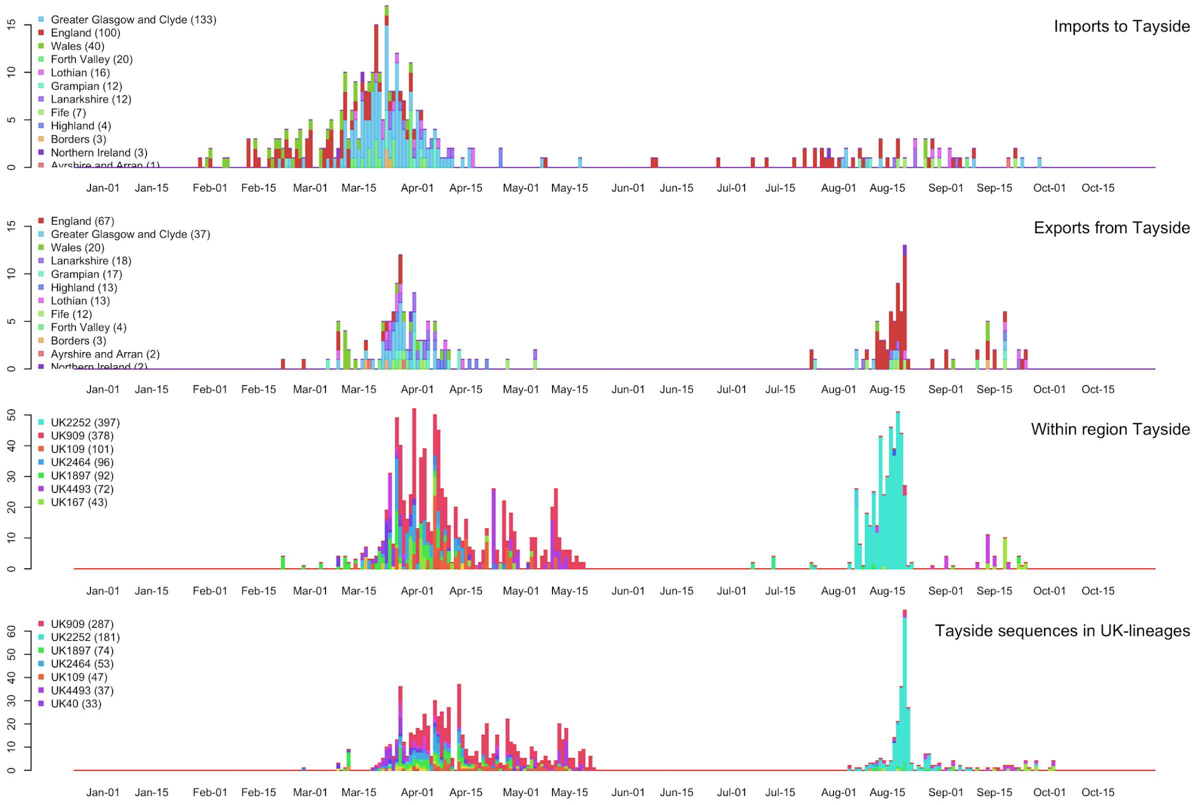


**Supplementary figure 7:** Timing of the imports and exports to Scottish NHS Health Boards with data to 2020-10-21. Results for Orkney, Shetland and the Western Isles are not included as there are too few sequences. The four plots for each NHS Health Board are as follows: top plot -- imports coloured by first NHS Health Board lineage detected in (rainbow colours) or England (red), Wales (green), Northern Ireland (purple). Note that the England (red) is a major source for both NHS Greater Glasgow and Clyde and Lothian in March; second row plot -- exports (same colouring as imports); third row plot -- within region transmissions coloured according to UK lineage number (other rainbow colours); bottom plot -- number of sequences per NHS Health Board coloured by UK lineage number (same as third row). Interactive plots can be found at [http://phylodynamics.lycett.roslin.ed.ac.uk:3838/RiseFallScotCOVID/](http://phylodynamics.lycett.roslin.ed.ac.uk:3838/RiseFallScotCOVID/)

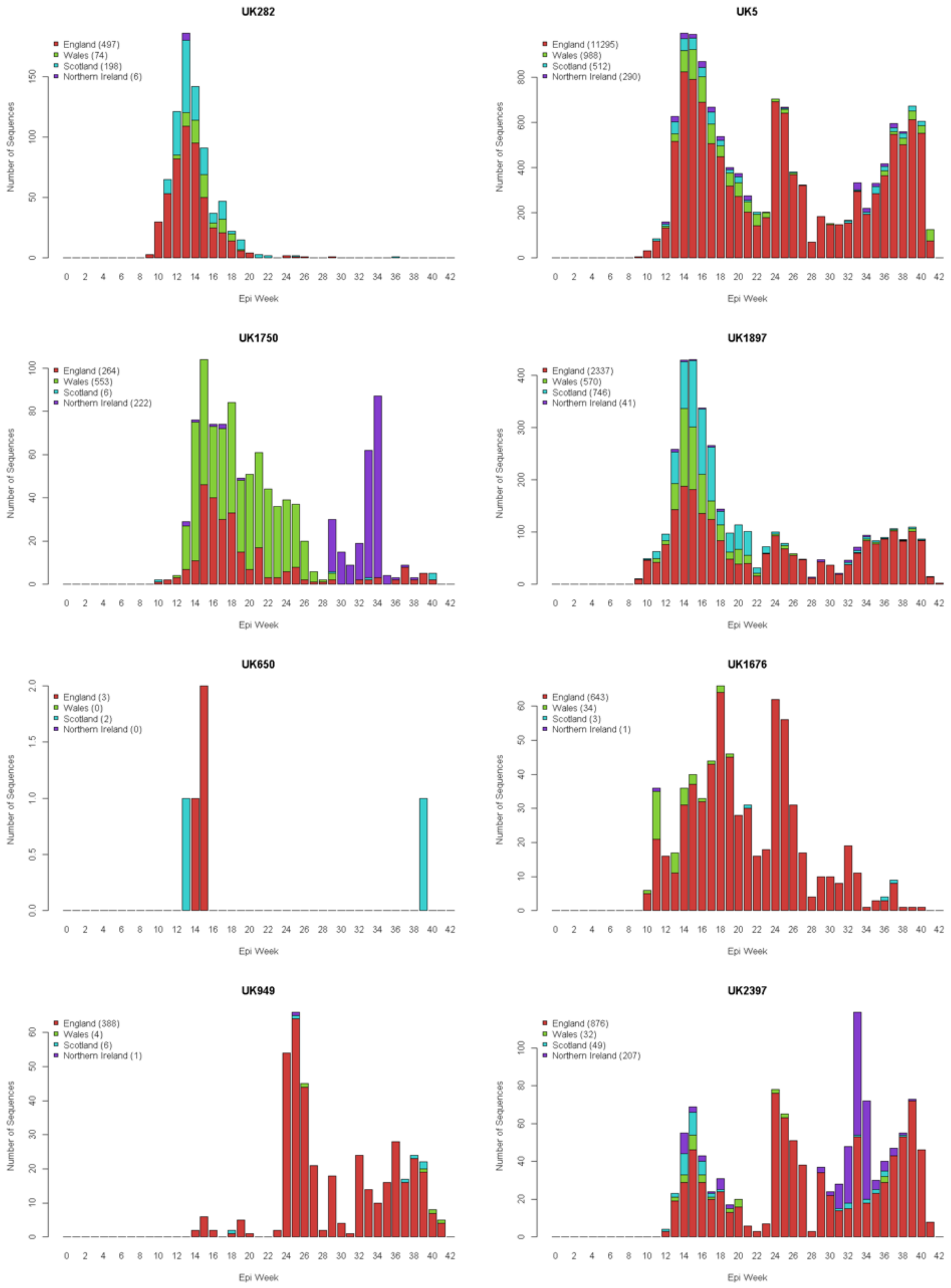


**Supplementary figure 8** (continued next page) Persisting lineages in Scotland -- number of sequences in England, Wales, Scotland and Northern Ireland with time for the selected ‘persisting in Scotland’ lineages, i.e., present in Scotland before 17th July and after 30th August.

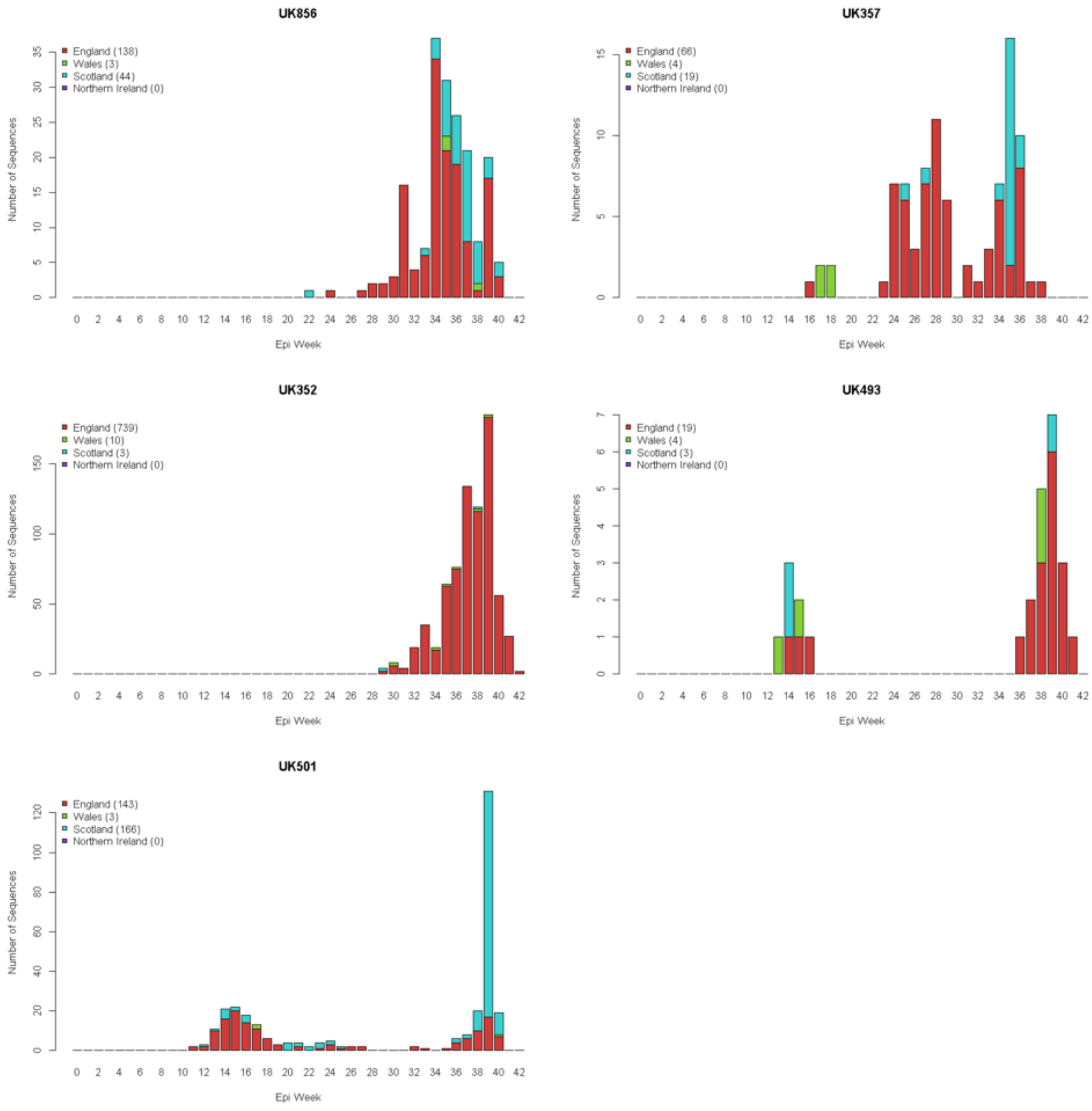


**Supplementary figure 8**. Persisting lineages in Scotland -- number of sequences in England, Wales, Scotland and Northern Ireland with time for the selected ‘persisting in Scotland’ lineages, i.e., present in Scotland before 17th July and after 30th August.

**Supplementary table 1**. Total size of UK lineages with sequences in Scotland both before epi-week 30 and after epi-week 36.

| Number of Sequences | UK650 | UK493 | UK357 | UK856 | UK501 | UK949 | UK1676 | UK352 | UK282 | UK1750 | UK2397 | UK1897 | UK5 |
|---|---|---|---|---|---|---|---|---|---|---|---|---|---|
| Total | 5 | 26 | 89 | 185 | 312 | 399 | 681 | 752 | 775 | 1045 | 1164 | 3694 | 13085 |
| England | 3 | 19 | 66 | 138 | 143 | 388 | 643 | 739 | 497 | 264 | 876 | 2337 | 11295 |
| Wales | 0 | 4 | 4 | 3 | 3 | 4 | 34 | 10 | 74 | 553 | 32 | 570 | 988 |
| Scotland | 2 | 3 | 19 | 44 | 166 | 6 | 3 | 3 | 198 | 6 | 49 | 746 | 512 |
| Northern Ireland | 0 | 0 | 0 | 0 | 0 | 1 | 1 | 0 | 6 | 222 | 207 | 41 | 290 |

**Supplementary table 2**. Summary of origin and date range for imported UK lineages to Scotland.

| Number of Imports | Before Mar 23 | Mar 23 – Jul 17 | Jul 17 – Aug 30 | After Aug 30 |
|---|---|---|---|---|
| All(inc Scot) | 317 | 153 | 50 | 9 |
| All(not Scot) | 285 | 131 | 46 | 9 |
| England | 111 | 66 | 28 | 8 |
| Wales | 8 | 7 | 1 | 0 |
| Scotland | 32 | 22 | 4 | 0 |
| Northern Ireland | 2 | 3 | 0 | 0 |
| Africa | 2 | 3 | 0 | 0 |
| Asia | 24 | 7 | 4 | 0 |
| Central America | 1 | 1 | 0 | 0 |
| Europe | 107 | 34 | 13 | 1 |
| North America | 15 | 6 | 0 | 0 |
| Oceania | 9 | 2 | 0 | 0 |
| South America | 6 | 2 | 0 | 0 |

**Supplementary table 3**. Relative contribution to sequences detected in Scotland by A| lineages originating in the summer (left), and B| lineages originating in the autumn (right).

| Summer Origin | Number of Lineages | Number of Scot. Seqs | Scot Contrib% | | Autumn Origin | Number of Lineages | Number of Scot. Seqs | Scot Contrib% |
|---|---|---|---|---|---|---|---|---|
| All(inc Scot) | 50 | 773 | 26.82 | | All(inc Scot) | 9 | 18 | 1.07 |
| All(not Scot) | 46 | 763 | 26.47 | | All(not Scot) | 9 | 18 | 1.07 |
| England | 28 | 255 | 8.85 | | England | 8 | 17 | 1.01 |
| Wales | 1 | 2 | 0.07 | | Wales | 0 | 0 | 0.00 |
| Scotland | 4 | 10 | 0.35 | | Scotland | 0 | 0 | 0.00 |
| Northern Ireland | 0 | 0 | 0.00 | | Northern Ireland | 0 | 0 | 0.00 |
| Africa | 0 | 0 | 0.00 | | Africa | 0 | 0 | 0.00 |
| Asia | 4 | 28 | 0.97 | | Asia | 0 | 0 | 0.00 |
| Central America | 0 | 0 | 0.00 | | Central America | 0 | 0 | 0.00 |
| Europe | 13 | 478 | 16.59 | | Europe | 1 | 1 | 0.06 |
| North America | 0 | 0 | 0.00 | | North America | 0 | 0 | 0.00 |
| Oceania | 0 | 0 | 0.00 | | Oceania | 0 | 0 | 0.00 |
| South America | 0 | 0 | 0.00 | | South America | 0 | 0 | 0.00 |

**Supplementary table 4.** – Details of summer lineage imports to Scotland (17th Jul to 30th August) -- lineages with >=3 Scottish sequences are listed (there are more with just 1 or 2 Scottish sequences).

| UKLineage | ImportEpiweek | ImportDate | Earliest case | England | Wales | Scotland | Northern Ireland | ScotContrib% |
|---|---|---|---|---|---|---|---|---|
| UK1909 | 30 | 24/07/2020 | Europe | 244 | 267 | 426 | 5 | 14.781 |
| UK1635 | 33 | 11/08/2020 | England | 1417 | 254 | 75 | 3 | 2.602 |
| UK847 | 30 | 21/07/2020 | England | 718 | 13 | 37 | 11 | 1.284 |
| UK1881 | 34 | 17/08/2020 | England | 507 | 7 | 26 | 0 | 0.902 |
| UK186 | 31 | 30/07/2020 | England | 113 | 24 | 21 | 0 | 0.729 |
| UK1119 | 34 | 18/08/2020 | Europe | 33 | 1 | 20 | 0 | 0.694 |
| UK924 | 31 | 31/07/2020 | England | 10 | 1 | 16 | 0 | 0.555 |
| UK1879 | 31 | 30/07/2020 | England | 192 | 56 | 13 | 4 | 0.451 |
| UK529 | 30 | 19/07/2020 | Asia | 1 | 0 | 13 | 0 | 0.451 |
| UK2837 | 33 | 15/08/2020 | England | 59 | 1 | 11 | 2 | 0.382 |
| UK918 | 31 | 27/07/2020 | Asia | 25 | 5 | 11 | 0 | 0.382 |
| UK1898 | 31 | 27/07/2020 | England | 48 | 4 | 10 | 5 | 0.347 |
| UK2402 | 29 | 18/07/2020 | Europe | 19 | 0 | 10 | 0 | 0.347 |
| UK2657 | 33 | 15/08/2020 | England | 29 | 0 | 7 | 0 | 0.243 |
| UK911 | 30 | 22/07/2020 | England | 2 | 0 | 6 | 0 | 0.208 |
| UK1084 | 34 | 20/08/2020 | England | 12 | 0 | 4 | 0 | 0.139 |
| UK1242 | 35 | 24/08/2020 | Scotland | 0 | 0 | 4 | 0 | 0.139 |
| UK1249 | 30 | 21/07/2020 | Europe | 118 | 5 | 4 | 3 | 0.139 |
| UK1656 | 32 | 03/08/2020 | England | 69 | 3 | 4 | 0 | 0.139 |
| UK1793 | 33 | 12/08/2020 | England | 24 | 0 | 4 | 1 | 0.139 |
| UK2079 | 35 | 25/08/2020 | Europe | 0 | 0 | 4 | 0 | 0.139 |
| UK2082 | 32 | 02/08/2020 | Europe | 0 | 0 | 4 | 0 | 0.139 |
| UK1538 | 31 | 31/07/2020 | Asia | 8 | 0 | 3 | 0 | 0.104 |
| UK2223 | 34 | 19/08/2020 | England | 28 | 0 | 3 | 0 | 0.104 |
| UK2591 | 33 | 12/08/2020 | Europe | 1 | 0 | 3 | 0 | 0.104 |

**Supplementary table 5.** Details of autumn lineage imports to Scotland from 30st August to 21st October (all listed).

| UKLineage | ImportEpiweek | ImportDate | Earliest case | England | Wales | Scotland | Northern Ireland | ScotContrib% |
|---|---|---|---|---|---|---|---|---|
| UK2369 | 38 | 16/09/2020 | England | 19 | 6 | 5 | 0 | 0.297 |
| UK855 | 37 | 06/09/2020 | England | 18 | 0 | 4 | 0 | 0.237 |
| UK1892 | 36 | 05/09/2020 | England | 73 | 1 | 2 | 1 | 0.119 |
| UK2592 | 36 | 02/09/2020 | England | 104 | 1 | 2 | 0 | 0.119 |
| UK1337 | 37 | 12/09/2020 | Europe | 1 | 0 | 1 | 0 | 0.059 |
| UK1464 | 36 | 03/09/2020 | England | 28 | 0 | 1 | 0 | 0.059 |
| UK1670 | 39 | 25/09/2020 | England | 173 | 3 | 1 | 0 | 0.059 |
| UK53 | 37 | 10/09/2020 | England | 22 | 2 | 1 | 1 | 0.059 |
| UK952 | 36 | 03/09/2020 | England | 58 | 1 | 1 | 3 | 0.059 |

**Supplementary table 6** -- GISAID acknowledgment file (see separate file).

**Appendix** -- COG-UK Authorship list (see separate file).
